# Associations of Combined Genetic and Lifestyle Risks with Incident Type 2 Diabetes in the UK Biobank

**DOI:** 10.1101/2024.12.16.24319115

**Authors:** Chi Zhao, Konstantinos Hatzikotoulas, Raji Balasubramanian, Elizabeth Bertone-Johnson, Na Cai, Lianyun Huang, Alicia Huerta-Chagoya, Margaret Janiczek, Chaoran Ma, Ravi Mandla, Amanda Paluch, Nigel W Rayner, Lorraine Southam, Susan R. Sturgeon, Ken Suzuki, Henry J Taylor, Nicole VanKim, Xianyong Yin, Chi Hyun Lee, Francis Collins, Cassandra N. Spracklen

## Abstract

**Background:** Type 2 diabetes (T2D) results from a complex interplay between genetic predisposition and lifestyle factors. Both genetic susceptibility and unhealthy lifestyle are known to be associated with elevated T2D risk. However, their combined effects on T2D risk are not well studied. We aimed to determine whether unhealthy modifiable health behaviors were associated with similar increases in the risk of incident T2D among individuals with different levels of genetic risk.

**Methods:** We performed a genetic risk score (GRS) by lifestyle interaction analysis within 332,251 non-diabetic individuals at baseline from the UK Biobank. Multi-ancestry GRS were calculated by summing the effects of 783 T2D-associated variants and ranked into tertiles. We used baseline self-reported data on smoking, BMI, physical activity level, and diet quality to categorize participants as having a healthy, intermediate, or unhealthy lifestyle. Cox proportional hazards regression models were used to generate adjusted hazards ratios (HR) of T2D risk and associated 95% confidence intervals (CI).

**Results:** During follow-up (median 13.6 years), 13,128 (4.0%) participants developed T2D. GRS (*P* < 0.001) and lifestyle classification (*P* < 0.001) were independently associated with increased risk for T2D. Compared with healthy lifestyle, unhealthy lifestyle was associated with increased T2D risk in all genetic risk strata, with adjusted HR ranging from 7.11 (low genetic risk) to 16.33 (high genetic risk).

**Conclusions:** High genetic risk and unhealthy lifestyle were the most significant contributors to the development of T2D. Individuals at all levels of genetic risk can greatly mitigate their risk for T2D through lifestyle modifications.

## Introduction

Type 2 diabetes (T2D) is among the leading causes of morbidity and mortality worldwide and has become one of the most challenging and concerning chronic diseases in public health. The prevalence of T2D has been steadily increasing over the past few decades: currently, > 28 million and > 500 million individuals have been diagnosed with T2D in the US and globally.^1,2^ Additionally, the prevalence of T2D is higher in certain populations in the US, ranging from 7.5% in non-Hispanic Whites to 14.7% in Native Americans/Alaska Natives, and both Hispanic Americans and non-Hispanic Blacks are almost twice as likely to have T2D as non-Hispanic Whites.^3^ The high prevalence and incidence of T2D results in a substantial disease burden, including treatment, reduced quality of life, disease complications, and premature death.

It is well-established that T2D is mainly caused by a complex interplay between genetic predisposition and lifestyle factors. Genome-wide association studies (GWAS) have identified more than 1,200 independent genetic variants associated with T2D, of which many appear to be related to insulin secretion and/or pancreatic β-cell development,^4,5^ and explain approximately 20-40% of the overall heritability for T2D.^6^ Lifestyle factors also play an important role in modulating T2D risk, and epidemiologic studies have identified increased T2D risk among individuals with a higher body mass index (BMI), low physical activity level, and unhealthy diet quality, as well as those who smoke.^7–10^ It is also known that lifestyle interventions can reduce the risk of development of T2D and improve cardiovascular health, especially among individuals at high risk of T2D.^11^

Several studies have attempted to examine the potential joint effects between genetic risk and overall behavioral and/or lifestyle factors on the risk for T2D.^12–19^ While most of the studies demonstrate the strongest T2D risk among those with the highest genetic risk and unhealthiest lifestyle factors, differences by biological sex were not often considered. Additionally, prior analyses were predominantly performed in East Asian-^14,15,17,18^ or European-ancestry^11–13,16,19^ individuals, limiting the generalizability to multi-ancestry and other non-European populations. Comparability across studies is further hindered by differences in behavioral and/or lifestyle factors considered as well as their classification and measurement. Finally, in terms of the GRS, the majority of the studies included fewer than 100 genetic variants in calculating the GRS,^11,14–19^ used effect size weights from a mismatched ancestry population,^15,18^ or used variants and effect size weights that were not independent of the study population,^12,17^ all of which limit the accuracy in measuring the genetic risk of T2D.

Therefore, the goal of this study was to determine whether unhealthy modifiable health behaviors, as determined by the American Heart Association, were associated with similar increases in the risk of incident T2D among individuals with different levels of genetic risk, utilizing the most up-to-date list of independent T2D-associated genetic variants, across all individuals in the UK Biobank.

## Methods

### Data Source

Details of the UK Biobank (UKB) study design and population have been described elsewhere.^20–22^ Briefly, the UKB is a population-based prospective cohort of > 500,000 participants designed to examine environmental, lifestyle, and genetic determinants of adult-onset diseases. Individuals aged 40-69 years old were recruited from 22 assessment centers throughout the United Kingdom from 2006-2010.^21^ At enrollment, participants provided extensive information on their demographics, health, and lifestyle through baseline questionnaires, interviews, and physical assessments. Blood samples were collected for genotyping; genotypes were then imputed to a merged UK10K and 1000 Genomes phase 3 panel.^22^ Follow-up of participants is ongoing through linked health records. The UKB study has approval from the North West Multi-center Research Ethics Committee, and all UKB participants provided written informed consent.

### Study Population

The study population was comprised of individuals from the UKB for which complete data were available for their genotypes, lifestyle factors at enrollment (BMI, smoking status, diet, and physical activity), and covariates. Participants were excluded if they: had a mismatch between their genetic and self-reported sex (n = 372), were missing genotypes for ≥ 7.5% of the included variants (n = 498), had a BMI at enrollment < 18.5 kg/m^2^ (n = 2,505), or had prevalent diabetes of any kind at enrollment (n = 15,250). To limit our analyses to those who developed incident T2D, we further excluded individuals who developed incident type 1 diabetes (T1D; ICD-10 E10), malnutrition-related diabetes (ICD-10 E12), or other specified diabetes (ICD-10 E13) (n = 699) during the follow-up period. Our final analytical sample size was n = 332,251 (Figure 1).

**Figure 1.**
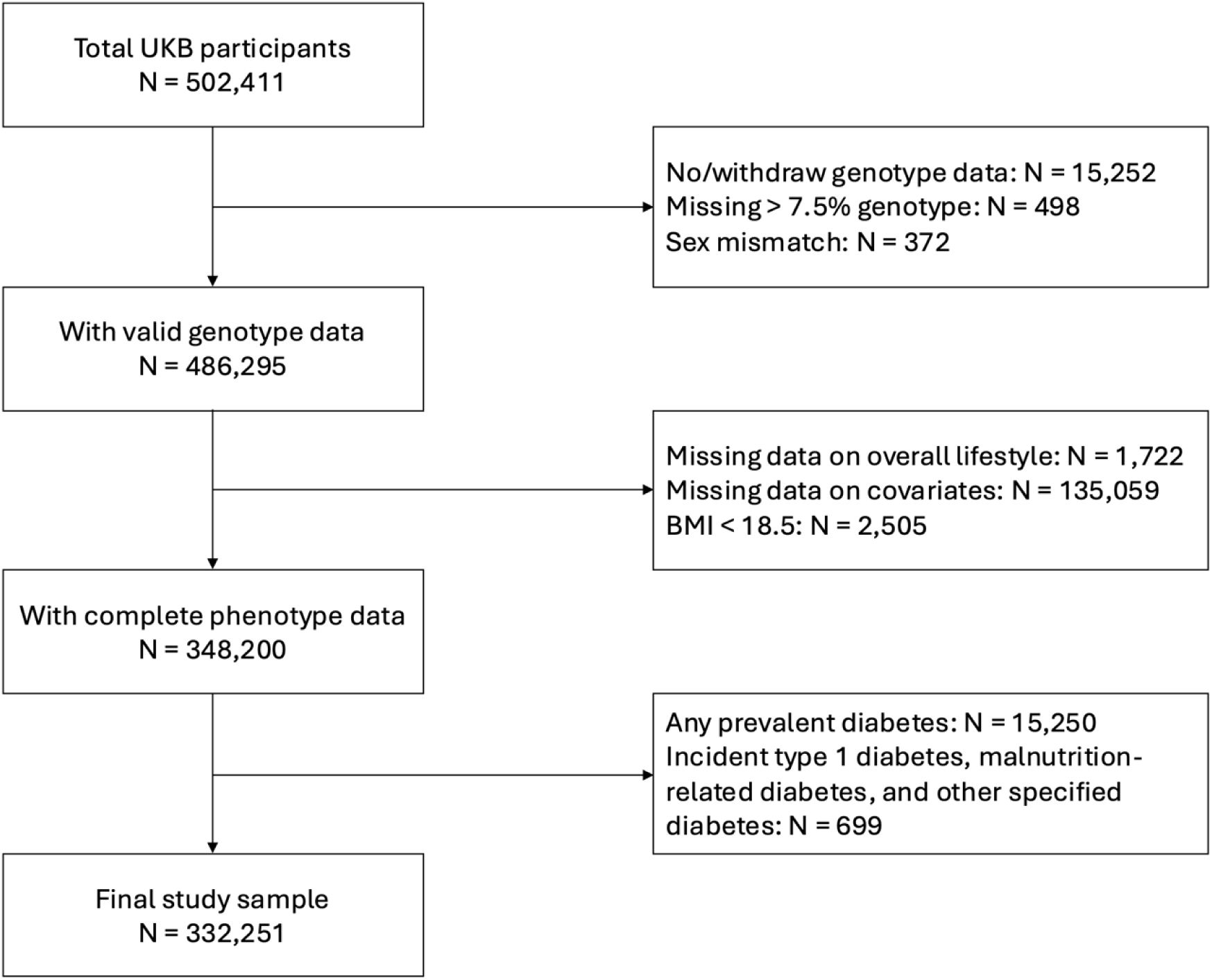
Flowchart of study population inclusion and exclusion.

### Genetic and Lifestyle Exposure groups

To estimate the genetic predisposition to T2D, genetic risk scores (GRS) were created following an additive model using 783 genome-wide significant variants identified from the most recent multi-ancestry GWAS meta-analysis (Table S1) after excluding results from the UKB, to avoid potential effect overestimation.^4^ The GRS were calculated using a weighted method in which, for each variant, the number of T2D risk-increasing alleles a person has is multiplied by the effect size estimate from the multi-ancestry fixed-effect meta-analysis; using PLINK 1.9,^23^ the products for each variant are then summed together for each individual into a continuous multi-ancestry GRS. GRS were divided into tertiles and categorized as low, moderate, and high genetic risk groups. We also generated ancestry-specific GRS for each individual using the effect size estimates from the global ancestry meta-analyses (African/African American, East Asian, European, South Asian) that most closely matched an individual’s self-reported ethnic background; individuals for which their self-reported ethnic background was “mixed”, “other”, or missing were omitted from the ancestry-specific analyses. To avoid potential effectmoverestimation, index variants and variant effect sizes were generated without inclusion of the UKB data in the meta-analyses. Genetic variants identified in the meta-analyses but missing in the UKB were excluded from the GRS.

We summarized an individual’s overall lifestyle into one of three categories (healthy, intermediate, and unhealthy) following the American Heart Association 2020 Strategic Impact Goal guidelines (termed ideal, intermediate, and poor, respectively) for smoking, BMI, and physical activity.^24^ Dietary priorities for cardiometabolic health were used to define a healthy or unhealthy category for diet quality.^25^ Definitions for healthy, intermediate, and unhealthy classifications for each of the lifestyle components can be found in the Supplementary Appendix (Table S2 and Table S3). Based on the categorization of the four lifestyle factors, we assigned participants to an overall lifestyle category: healthy (having at least 3 healthy lifestyle factors), unhealthy (having at least 3 unhealthy lifestyle factors), or intermediate (all other combinations).

### Ascertainment of Incident T2D Outcomes

Incident cases of T2D within the UKB were identified using the first occurrences data. The first occurrences dataset indicates the first occurrence of any disease (mapped to ICD-10 diagnosis codes) from primary care, hospital inpatient, death register, and self-reported data.^21^ For this study, we used the first record of ICD-10 diagnosis code E11 (type 2 diabetes mellitus) and the corresponding date to define incident cases.

### Statistical Analysis

Descriptive statistics for participants were generated using baseline data and compared between censored observations and incident T2D cases using t-tests for continuous variables and chi-squared test for categorical variables. For this analysis, participants were followed from enrollment until diagnosis of T2D, death, lost to follow-up, or censoring date (by the time of analysis, the censoring date is 2022-10-31 for individuals in England, 2021-07-31 for individuals in Scotland, and 2018-02-28 for individuals in Wales), whichever came first. Several multivariable Cox regression models were used to test both the independent (both genetic risk and lifestyle as predictor variables) and joint associations of genetic risk and lifestyle groups with incident T2D; hazards ratios (HR) and associated 95% confidence intervals (95% CI) were calculated using individuals with low GRS and healthy lifestyle as the reference group. Additionally, we tested the independent association of genetic risk and all the individual lifestyle factors. The proportional hazards assumption was tested based on visualization of the survival probabilities over time and the scaled Schoenfeld residuals; the assumptions were met (Supplementary Figures S1-S10). Adjusted models included the following covariates: age at baseline, biological sex, years in education,^26^ Townsend Deprivation Index (TDI),^27^ income, and the first 16 genetic principal components (to adjust for population subtructure).^23^ Sex-stratified analyses were also conducted including the same covariates except for biological sex. We also tested for statistical interaction between the GRS and lifestyle factors. Finally, we calculated the population attributable fraction (PAF), to evaluate the proportion of incident T2D that would have been prevented if participants with intermediate or unhealthy lifestyle had been in the healthy category. All analyses were additionally conducted using the ancestry-specific GRS that most closely matched the self-reported ethnic background. All statistical tests were two-sided, and P-values < 0.05 were considered statistically significant. All analyses were conducted using R 4.3.0.

## Results

Baseline characteristics of study participants can be found in Table 1. Overall, the mean (SD) age was 55.19 (8.06) years, and 177,869 (54%) were female. During a median follow-up of 13.56 (IQR: 12.74-14.25) years, 13,128 (4%) participants developed incident T2D during a median time to onset of 7.98 years (IQR: 4.92, 10.82), with higher incidence rates among the high GRS tertile and unhealthy lifestyle classifications (0.75 per 1,000 person-years for those with the lowest GRS and healthy lifestyle to 12.53 per 1,000 person-year for those with the highest GRS and unhealthy lifestyle; Table 2; Supplementary Figure S11). At baseline, participants who later developed T2D were older, more likely to be male, have fewer years of education, lower income, and more severe social deprivation than those who did not develop T2D during the follow-up period. Individuals who developed incident T2D also had a significantly higher mean GRS and were more likely classified in the moderate or high GRS tertile. Finally, compared to the censored observations, incident T2D cases were more likely to have a higher BMI, lower physical activity level, lower diet quality, and to be a current smoker, which resulted in a significantly higher proportion of incident T2D cases also falling into the “unhealthy” lifestyle category.

**Table 1.**
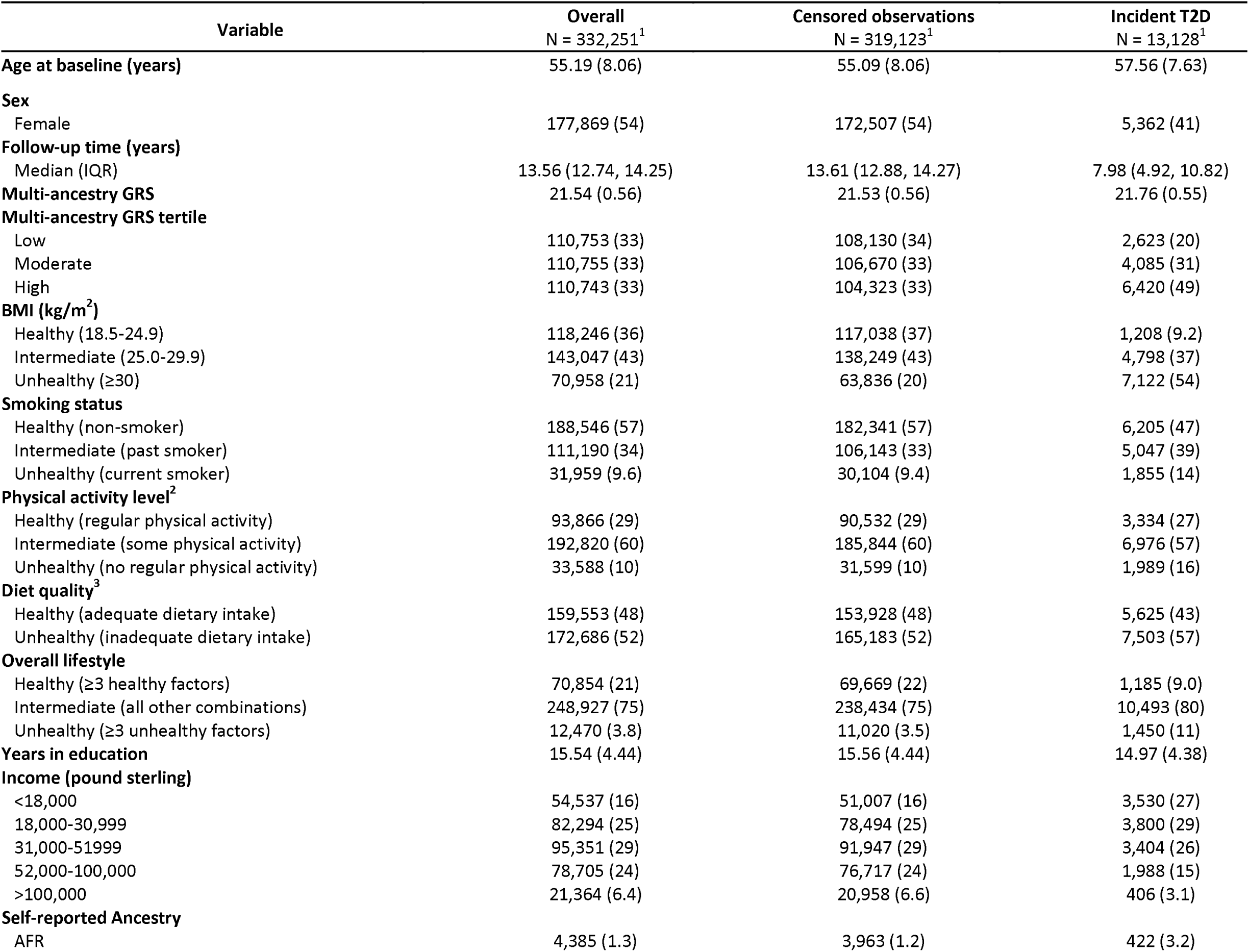
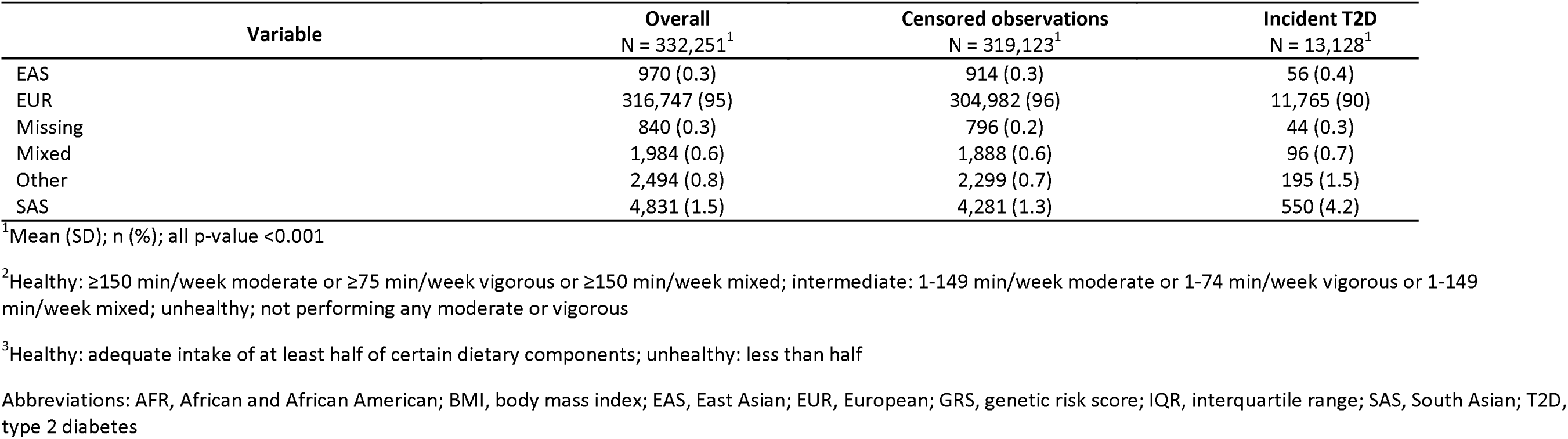
Baseline characteristics of 332,251 participants from the UK Biobank.

**Table 2.** Incidence rates of T2D by the category of combined genetic and lifestyle risk.

| Multi-ancestry GRS<br>tertile | Lifestyle | N total (%) | N case (%<sup>1</sup>) | Person-year | Incidence<br>rate<sup>2</sup> |
| --- | --- | --- | --- | --- | --- |
| Low | Healthy | 24,408 (7.4) | 245 (1.0) | 325,304.3 | 0.75 |
|  | Intermediate | 82,402 (24.8) | 2,082 (2.5) | 1,083,869.8 | 1.92 |
|  | Unhealthy | 3,943 (1.2) | 296 (7.5) | 50,042.0 | 5.92 |
| Moderate | Healthy | 23,373 (7.0) | 347 (1.5) | 311,018.5 | 1.12 |
|  | Intermediate | 83,224 (25.1) | 3,250 (3.9) | 1,088,448.2 | 2.99 |
|  | Unhealthy | 4,158 (1.3) | 488 (11.7) | 51,563.5 | 9.46 |
| High | Healthy | 23,073 (6.9) | 593 (2.6) | 305,394.7 | 1.94 |
|  | Intermediate | 83,301 (25.1) | 5,161 (6.2) | 1,078,391.9 | 4.79 |
|  | Unhealthy | 4,369 (1.3) | 666 (15.2) | 53,161.2 | 12.53 |

<sup>1</sup>Cumulative incidence
<sup>2</sup>The incidence rate is provided per 1,000 person-years.

GRS tertiles (moderate, HR: 1.59, 95% CI 1.52-1.67; high, HR: 2.58, 95%CI: 2.47-2.70) and overall lifestyle categories (intermediate, HR: 2.38, 95% CI 2.24-2.52; unhealthy, HR: 6.83, 95% CI 6.32-7.38) were independently associated with T2D risk (Supplementary Table S4). For the standardized GRS, a 1-SD increase was associated with a 53% increased risk of T2D (HR: 1.53, 95% CI: 1.50-1.55; Supplementary Table S5). Results were similar for individual lifestyle factors and in models stratified by sex (Supplementary Tables S6-S7); BMI had the strongest independent association with incident T2D (intermediate, HR: 2.81, 95% CI 2.63-3.00; unhealthy, HR: 8.84, 95% CI 8.29-9.42).

Across all GRS tertiles, individuals classified as having a unhealthy lifestyle were at substantially increased risk for T2D compared to those classified as having an healthy lifestyle, with HR ranging from 7.11 (low GRS tertile) to 16.33 (high GRS tertile; Figure 2). Compared to those in the healthy lifestyle group, individuals in the intermediate lifestyle group were also at increased risk for T2D, with a 2-, 4-, and 6- fold increased risk for those in the low, moderate, and high GRS tertile. When focusing within a single genetic risk tertile (e.g., low GRS), individuals in the unhealthy lifestyle category were at a 6- to 8-fold increased risk of T2D compared to the healthy lifestyle. Results were similar in sex-stratified analyses (Supplemental Table S8). While the effect estimates were slightly stronger in females than in males, the difference was not statistically significant (*P* = 0.39). We did not detect a significant interaction between the GRS tertiles and lifestyle classification, however we did detect a significant interaction when considering the standardized (continuous) GRS and lifestyle classifications (GRS*intermediate lifestyle, p<0.20; GRS*unhealthy lifestyle, p=0.004; Supplementary Tables S9-S10). Lastly, results showed similar trends across the self-reported ancestry groups but were generally underpowered in many GRS tertile/lifestyle category combinations for analyses of non-European-ancestry individuals (Supplemental Table S11).

When calculating the ancestry-specific GRS using the same set of genetic variants, but with the ancestry-specific weights, we found similar trends in the associations between combined GRS and lifestyle risk with incident T2D (Supplemental Tables S12-16; Supplementary Figures S12-14).

To evaluate the proportion of incident T2D that would have been prevented if subjects with intermediate or unhealthy lifestyle (also considered “non-healthy”) had instead been in the healthy category, we calculated the population attributable fraction (PAF; Supplementary Table S17). Regardless of the GRS, more than 55% of incident T2D cases in the UKB would have been prevented if all individuals in the “non-healthy” lifestyle categories would have been in the healthy lifestyle category (Year 1: 95% CI 0.53-0.58). The PAF proportions were consistent across each time point during the 15 years of follow-up.

## Discussion

In this large population-based prospective cohort study with over 332,000 multi-ancestry participants from the UK Biobank, both high GRS and unhealthy lifestyle were independently associated with increased risk of T2D. Across and within different GRS tertiles, adherence to an intermediate or unhealthy lifestyle was associated with substantially increased risk of T2D compared to an healthy lifestyle. Overall, while our analyses support the notion that while genetics play a large role in the risk for developing T2D and T2D etiology, lifestyle factors play a substantially larger role, particularly BMI. Further, we demonstrated that individuals with any level of genetic risk could greatly reduce their risk for T2D through modifiable healthy lifestyles.

To our knowledge, this study is the first to test the effect of combined lifestyle factors in different genetic risk level for T2D based on nearly 800 genetic variants and the first to consider both a multi-ancestry and ancestry-matched GRS. Consistent with findings from prior studies^11–19^, we found high GRS and unhealthy lifestyle factors were independently and jointly associated with increased risk of developing T2D. However, there is wide variability in effect sizes across the prior studies, most likely due to fundamental differences in study design and methodology, including sample size, T2D GRS composition and calculation, and consideration of behavioral and lifestyle factors. Most similar to this study, Said, et al. previously used the UKB study population to examine the combined effects of genetic and lifestyle risk of T2D.^11^ Among 322,014 individuals, the study also found strong effects of unhealthy lifestyle across different GR tertiles, with adjusted HR ranging from 10.82 to 15.46 in sex-combined analyses. While both studies used a similar approach to categorize lifestyle factors based on the American Heart Association guidelines, the prior study included only the European-ancestry individuals within the UKB, had a less-restrictive definition for incident T2D that likely resulted in outcome misclassification, and calculated the GRS based on only 38 variants. Our study included a much more comprehensive measure of GRS (783 variants), excluded individuals from the analyses if they did not have confirmed T2D, and did not exclude individuals based on genetic ancestry.

Based on the results presented, it is clear that individuals who have either moderate or higher genetic risk for T2D with intermediate or unhealthy lifestyle are at substantially increased risk for T2D. These findings indicate the strong potential benefits of adherence to multiple healthy lifestyle factors to mitigate disease risk, regardless of genetic risk. In fact, our analysis suggests that 55% of incident T2D risk in the population that could be theoretically eliminated if individuals with non-healthy lifestyle were to be shifted to be having healthy lifestyle, highlighting the potential impact of shifting individuals from the non-healthy to the healthy lifestyle category. Although challenges remain in communicating individual genetic risk information to patients that is understandable and interpretable by the general population, knowledge of the strong impact a healthy lifestyle can have to mitigate genetic or familial risk for T2D may motivate patients to change behaviors

### Strengths and Limitations

To our knowledge, this is the first study to investigate the associations of combined genetic and lifestyle risk of T2D using the most up-to-date set of T2D-associated variants.^4^ Major strengths of the study were the prospective cohort design, large sample size, and comprehensive measure of genetic risk. The list of T2D-associated variants and their effect sizes used for our GRS calculation were determined independently of the UKB study population. Our study also utilized all individuals in the UKB, regardless of self-reported ancestry, and used both combined and ancestry-specific genetic effect sizes when calculating the GRS, which improves the external validity of our findings. Further, we classified all of the lifestyle factors into healthy, intermediate, and unhealthy based on guidelines from the American Heart Association,^24^ which allows for a more direct clinical interpretation of our results.

There are also several limitations. Measurements for all lifestyle factors were obtained at study entry, of which three were based on self-reported data. Because they are all potentially time-varying covariates, misclassification of exposure is possible. However, due to the prospective design of the UKB, any misclassification would be nondifferential and would bias the result towards the null, resulting in an underestimation of the true association. Second, incident T2D cases were identified using the first occurrence data in the UK Biobank, which includes self-reported outcomes. Further, the suspected rate of undiagnosed T2D in the UK is estimated to be around 2%.^28^ Thus, misclassification of some T2D cases as non-cases is possible; however, we would expect this bias the results toward the null. The genetic variants used in the GRS calculation may also have pleiotropic effects on lifestyle factors, including BMI. Although our study included individuals with diverse ethnic backgrounds, the generalizability of the findings remains somewhat limited due to the predominance of European participants in the UKB.

In conclusion, both genetic risk and lifestyle were independently associated with elevated T2D risk, but individuals with the unhealthiest lifestyle were at the highest risk for incident disease. Comprehensive and multifactorial lifestyle modifications should be encouraged in individuals at all levels of genetic risk to greatly mitigate their risk of developing T2D, though individuals at the highest levels of genetic risk will gain the most benefit. Further studies investigating the joint effects of lifestyle changes over time and their interplay with genetics for the T2D risk is warranted.

## Data Availability

All data produced in the present work are contained in the manuscript

## Acknowledgments

The present study was conducted under application #81009 of the UK Biobank resource.

## Funding

CNS and CZ were supported by the NIH (R01DK118011) and the American Diabetes Association (11-22-JDFPM-06). CNS was further supported by the NIH (R01DK136671). EBJ was supported by the NIH (R34MH118396 and K01DK123193) and the Commonwealth of Massachsutts (INTF4107H78241733154). NVK was support by the NIH (K01DK123193). AH-C is supported by the American Diabetes Association grant 11-23-PDF-35.

---

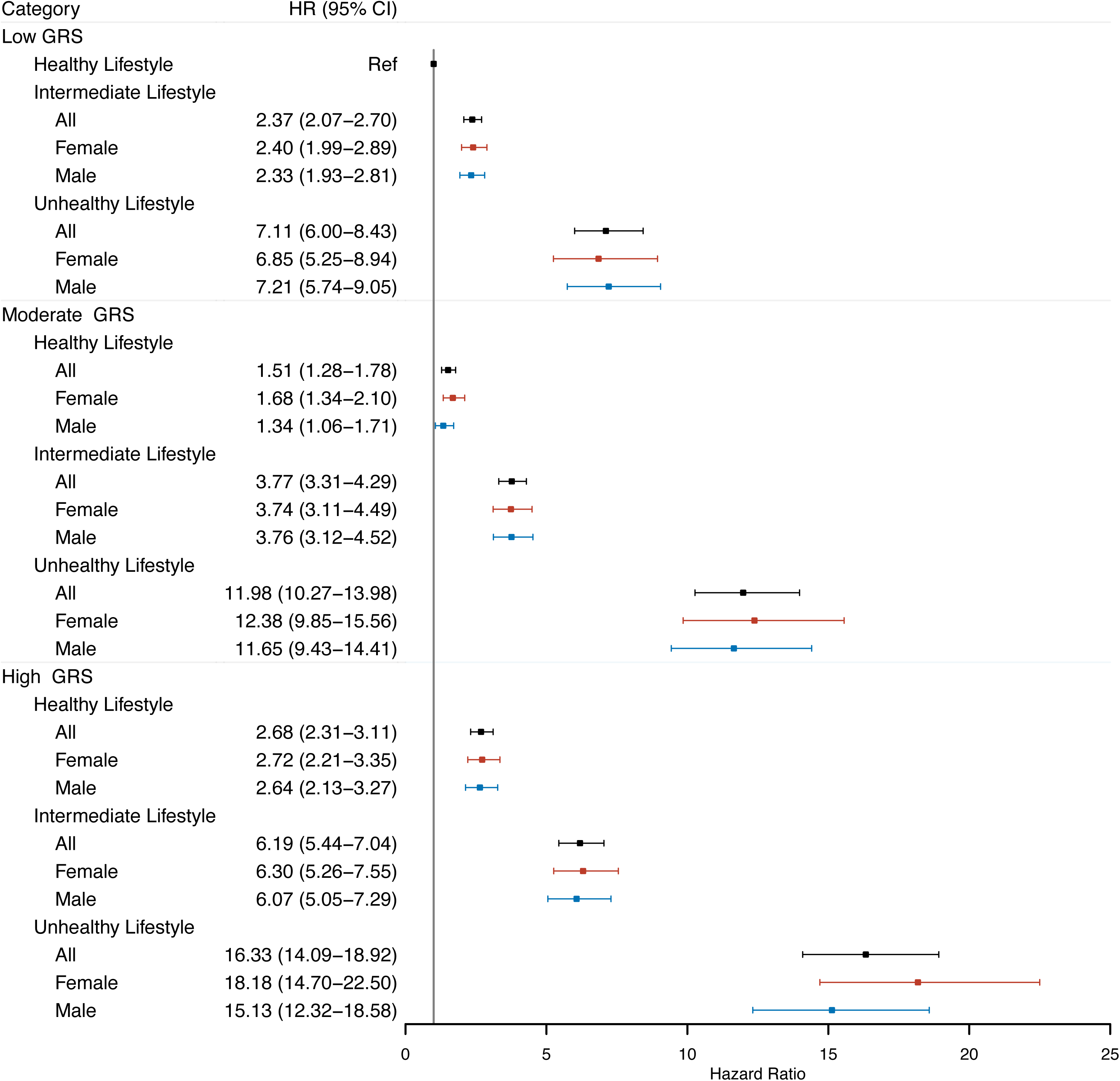

## Notes

### Competing Interest Statement

The authors have declared no competing interest.

### Author Declarations

The UKB study has approval from the North West Multi-center Research Ethics Committee, and all UKB participants provided written informed consent.

