## Supplemental Appendix for "Associations of Combined Genetic and Lifestyle Risks with Incident Type 2 Diabetes in the UK Biobank"

This appendix has been provided by the authors to give readers additional information about the work.

Associations of Combined Genetic and Lifestyle Risks with Incident Type 2 Diabetes in the UK Biobank

Chi Zhao^1^, Konstantinos Hatzikotoulas^2^, Raji Balasubramanian, ScD^1^, Elizabeth Bertone-Johnson, ScD^3^, Na Cai, PhD^4,5,6^, Lianyun Huang^4,5,6^, Alicia Huerta-Chagoya^7^, Margaret Janiczek^1^, Chaoran Ma, MD, PhD^3^, Ravi Mandla^7,8,9^, Amanda Paluch, PhD^10^, Nigel W Rayner^2^, Lorraine Southam^2^, Susan R. Sturgeon, DrPH^1^, Ken Suzuki, PhD, MD^12,13^, Henry J Taylor^14,15,16^, Nicole VanKim, PhD^1^, Xianyong Yin^17,18^, Chi Hyun Lee, PhD^19^, Francis Collins, PhD, MD^20^, Cassandra N. Spracklen, PhD^1^

^1^Department of Biostatistics and Epidemiology, University of Massachusetts Amherst, Amherst, MA, USA

^2^Institute of Translational Genomics, Helmholtz Zentrum München, German Research Center for Environmental Health, Neuherberg, Germany

^3^Department of Nutrition, University of Massachusetts Amherst, Amherst, MA, USA

^4^Helmholtz Pioneer Campus, Helmholtz Munich, Neuherberg, Germany

^5^Computational Health Centre, Helmholtz Munich, Neuherberg, Germany

^6^School of Medicine and Health, Technical University of Munich, Munich, Germany

^7^Programs in Metabolism and Medical and Population Genetics, Broad Institute of Harvard and MIT, Cambridge, MA, USA

^8^Diabetes Unit and Center for Genomic Medicine, Massachusetts General Hospital, Boston, MA, USA

^9^Graduate Program in Genomics and Computational Biology, University of Pennsylvania, Philadelphia, PA, USA

^10^Department of Kinesiology, Institute for Applied Life Sciences, University of Massachusetts Amherst, Amherst, MA, USA

^12^Department of Diabetes and Metabolic Diseases, Graduate School of Medicine, University of Tokyo, Tokyo, Japan

^13^Department of Statistical Genetics, Osaka University Graduate School of Medicine, Suita, Japan

^14^Center for Precision Health Research, National Human Genome Research Institute, National Institutes of Health, Bethesda, MD, USA

^15^British Heart Foundation Cardiovascular Epidemiology Unit, Department of Public Health and Primary Care, University of Cambridge, Cambridge, UK

^16^Heart and Lung Research Institute, University of Cambridge, Cambridge, UK

^17^Department of Epidemiology, School of Public Health, Nanjing Medical University, Nanjing, China

^18^Department of Biostatistics and Center for Statistical Genetics, University of Michigan, Ann Arbor, MI, USA

^19^Department of Applied Statistics, Yonsei University, Seoul, South Korea

^20^National Human Genome Research Institute, National Institutes of Health, Bethesda, MD, USA

^*^Corresponding author:

Cassandra N. Spracklen

Department of Biostatistics and Epidemiology, 715 North Pleasant Street, University of Massachusetts Amherst, Amherst, MA, USA

Table of Contents

Table S1. Genetic variants used to build the genetic risk scores for T2D (clumped)	#

Table S2. Lifestyle factor definitions in the UK Biobank Study	#

Table S3. Dietary quality components definitions in the UK Biobank Study	#

Table S4. Independent associations of GRS tertiles (multi-ancestry) and the three-level
lifestyle category with incident T2D risk	#

Table S5. Independent associations of continuous GRS (multi-ancestry) and the three-level lifestyle	category with incident T2D risk	#

Table S6. Independent associations of GRS tertile (multi-ancestry) and individual lifestyle factors with
 incident T2D risk	#

Table S7. Independent associations of continuous GRS (multi-ancestry) and individual lifestyle factors
 with incident T2D risk	#

Table S8. Associations of continuous GRS (multi-ancestry) and the three-level lifestyle category with
incident T2D risk, including the multiplicative interaction between the GRS and lifestyle category	#

Table S9. Associations of GRS tertile (multi-ancestry) and the three-level lifestyle category with incident
 T2D risk, including the multiplicative interaction between the GRS tertiles and lifestyle category	#

Table S10. Association of combined genetic (multi-ancestry) and lifestyle risk and T2D stratified by
ancestry	#

Table S11. Population attributable fraction	#

Table S12. Association of combined genetic (ancestry-specific) and lifestyle risk and T2D	#

Table S13. Independent associations of GRS tertile (ancestry-specific) and individual lifestyle factors
with incident T2D risk	#

Table S14. Independent associations of GRS (ancestry-specific) and individual lifestyle factors with
incident T2D risk	#

Table S15. Independent associations of GRS tertiles (ancestry-specific) and the three-level lifestyle
category with incident T2D risk	#

Table S16. Independent associations of continuous GRS (ancestry-specific) and the three-level lifestyle category with incident T2D risk	#

Table S17. Association of combined genetic (ancestry-specific) and lifestyle risk and T2D stratified by ancestry	#

Figure S1. Multi-ancestry GRS by T2D	#

Figure S2. Ancestry-specific GRS by T2D	#

Figure S3. Correlation between multi-ancestry and ancestry-specific GRS	#

Figure S4. Survival curves for multi-ancestry GR category	#

Figure S5. Survival curves for ancestry-specific GR category	#

Figure S6. Survival curves for lifestyle classification	#

Figure S7. Survival curves for combined genetic (multi-ancestry) and lifestyle risk	#

Figure S8. Survival curves for combined genetic (ancestry-specific) and lifestyle risk	#

Figure S9. Graphical test of proportional hazard (PH) assumption for multi-ancestry GRS	#

Figure S10. Graphical test of PH assumption for multi-ancestry GR	#

Figure S11. Graphical test of PH assumption for lifestyle classification	#

Figure S12. Graphical test of PH assumption for combined genetic (multi-ancestry) and
 lifestyle risk	#

Figure S13. Graphical test of PH assumption for combined genetic (ancestry-specific) and
 lifestyle risk	#

Figure S14. Association of combined genetic (ancestry-specific) and lifestyle risk with T2D	#

**Table S1. Genetic variants used to build the genetic risk scores for T2D**

Please see the TableS1.xlsx file. **Table S2. Lifestyle factor definitions in the UK Biobank Study**

| **Lifestyle factors (Field ID)** | **Healthy** | **Intermediate** | **Unhealthy** |
| --- | --- | --- | --- |
| Smoking (20116) | nonsmokers | past smokers | current smokers |
| BMI (21001) | 18.5-24.9 kg/m^2^ | 25-29.9 kg/m^2^ | ≥30 kg/m^2^ |
| Leisure-time Physical activity^1^ (884, 894, 904, 914) | ≥150 min/week moderate or  ≥75 min/week vigorous or  ≥150 min/week mixed | 1-149 min/week moderate or  1-74 min/week vigorous or  1-149 min/week mixed | not performing any moderate or vigorous |
| Diet (see Table S4) | adequate intake of at least half of certain dietary components | N/A | less than half |
| **Overall classification** | ≥ 3 ideal lifestyle factors | all other combinations | ≥ 3 poor lifestyle factors |

*^1^Leisure-time physical activity was defined based on IPAQ (International Physical Activity Questionnaire) guidelines*

Table S3. Dietary quality components definitions in the UK Biobank Study

| **Diet component** | **Intake goal** | **Field IDs** | **Amount per serving** |
| --- | --- | --- | --- |
| Fruit | 3 servings/day | 1309 (pieces fresh fruit/day) 1319 (pieces dried fruit/day) | 1309 – 1 piece 1319 – 5 pieces |
| Vegetable | 3 servings/day | 1289 (tablespoons cooked vegetables/day) 1299 (salad/raw vegetables/day) | 3 heaped tablespoons |
| Whole grains | 3 servings/day | 1438, 1448 (whole meal/wholegrain bread slices/week) 1458, 1468 (bran/oat/muesli cereal bowls/week) | 1438/1448 – 1 slice/day 1458/1468 – 1 bowl/day |
| (Shell) fish | ≥2 servings/week | 1329 (oily fish/week) 1339 (non-oily fish/week) | Once/week |
| Dairy | 2 servings/day | 1408 (cheese/week) 1418 (milk type) | 1408 – 1 piece/day 1418 – 1 glass/day if consumption of any type of milk |
| Vegetable oils | 2 servings/day | 1428 (Flora Pro-Active/Benecol spread) 2654 (Flora Pro-Active/Benecol, soft margarine -, olive oil based -, polyunsaturated/sunflower oil based -, other low/reduced fat spread) 1438 (bread slices/week) | 1 serving/day if in combination with eating at least 2 slices of bread (ID 1438) |
| Refined grains | ≤2 servings/day | 1438, 1448 (white, brown, other bread slices/week) 1458, 1468 (biscuit, other cereals/week) | 1438/1448 – 1 slice/day 1458/1468 – 1 bowl/day |
| Processed meats | ≤1 serving/week | 1349 (processed meat/week or daily) 3680 (age when last ate meat) | 1349 – 1 piece/day 3680 – 0 pieces/day if indicated having never eaten meat |
| Unprocessed meats | ≤2 serving/week | 1359 (poultry/week or day) 1369 (beef/week or day) 1379 (lamb or mutton/week or day) 1389 (pork/week or day) 3680 (age when last ate meat) | 1359-1389 – once/week 3680 – 0 pieces/day if indicated having never eaten meat |
| Sugar-sweetened beverages | Don’t drink | 6144 (never consumes drinks containing sugar) | Only 0 servings were possible here. |

Table S4. Independent associations of GRS tertiles (multi-ancestry) and the three-level lifestyle category with incident T2D risk

|  | **Sex-combined** | | | | **Female** | | | **Male** | | |
| --- | --- | --- | --- | --- | --- | --- | --- | --- | --- | --- |
| **Characteristic** | **HR** | | **95% CI** | **p-value** | **HR** | **95% CI** | **p-value** | **HR** | **95% CI** | **p-value** |
| Multi-ancestry GRS tertile | |  |  |  |  |  |  |  |  |  |
| Low | | — | — |  | — | — |  | — | — |  |
| Moderate | | 1.59 | 1.52, 1.67 | <0.001 | 1.60 | 1.48, 1.73 | <0.001 | 1.59 | 1.49, 1.70 | <0.001 |
| High | | 2.58 | 2.47, 2.70 | <0.001 | 2.64 | 2.46, 2.84 | <0.001 | 2.54 | 2.39, 2.69 | <0.001 |
| Lifestyle classification | |  |  |  |  |  |  |  |  |  |
| Healthy | | — | — |  | — | — |  | — | — |  |
| Intermediate | | 2.38 | 2.24, 2.52 | <0.001 | 2.30 | 2.12, 2.51 | <0.001 | 2.44 | 2.24, 2.67 | <0.001 |
| Unhealthy | | 6.83 | 6.32, 7.38 | <0.001 | 6.93 | 6.17, 7.79 | <0.001 | 6.84 | 6.15, 7.61 | <0.001 |
| *Results from Cox proportional hazards model including the multi-ancestry GRS tertiles and the three-level lifestyle classification variable; model is further adjusted for age at enrollment, sex (sex-combined only), years in education, household income, Townsend Deprivation Index, and the first 16 genotype PC’s.*  *Abbreviations: CI, confidence interval; GRS, genetic risk score; HR, hazard ratio; T2D, type 2 diabetes* | | | | | | | | | | |

Table S5. Independent associations of continuous GRS (multi-ancestry) and the three-level lifestyle category with incident T2D risk

|  | | **Sex-combined** | | | **Female** | | | **Male** | | |
| --- | --- | --- | --- | --- | --- | --- | --- | --- | --- | --- |
| **Characteristic** | **HR** | | **95% CI** | **p-value** | **HR** | **95% CI** | **p-value** | **HR** | **95% CI** | **p-value** |
| Multi-ancestry GRS | 1.53 | | 1.50, 1.55 | <0.001 | 1.54 | 1.50, 1.58 | <0.001 | 1.52 | 1.48, 1.55 | <0.001 |
| Lifestyle classification |  | |  |  |  |  |  |  |  |  |
| Healthy | — | | — |  | — | — |  | — | — |  |
| Intermediate | 2.37 | | 2.23, 2.52 | <0.001 | 2.30 | 2.11, 2.50 | <0.001 | 2.44 | 2.24, 2.66 | <0.001 |
| Unhealthy | 6.82 | | 6.31, 7.37 | <0.001 | 6.92 | 6.16, 7.77 | <0.001 | 6.84 | 6.14, 7.61 | <0.001 |
| *Results from Cox proportional hazards model including the continuous GRS value and the three-level lifestyle classification variable; model is further adjusted for age at enrollment, sex (sex-combined only), years in education, household income, Townsend Deprivation Index, and the first 16 genotype PC’s. The association tested was based on 1-standard deviation increase in the multi-ancestry GRS.*  *Abbreviations: CI, confidence interval; GRS, genetic risk score; HR, hazard ratio; T2D, type 2 diabetes* | | | | | | | | | | |

**Table S6. Independent associations of GRS tertile (multi-ancestry) and individual lifestyle factors with incident T2D risk**

|  | **Sex-combined** | | | **Female** | | | **Male** | | |
| --- | --- | --- | --- | --- | --- | --- | --- | --- | --- |
| **Characteristic** | **HR** | **95% CI** | **p-value** | **HR** | **95% CI** | **p-value** | **HR** | **95% CI** | **p-value** |
| Multi-ancestry GRS tertile |  |  |  |  |  |  |  |  |  |
| Low | — | — |  | — | — |  | — | — |  |
| Moderate | 1.58 | 1.50, 1.66 | <0.001 | 1.57 | 1.45, 1.70 | <0.001 | 1.58 | 1.48, 1.68 | <0.001 |
| High | 2.52 | 2.40, 2.64 | <0.001 | 2.57 | 2.38, 2.77 | <0.001 | 2.49 | 2.34, 2.64 | <0.001 |
| BMI |  |  |  |  |  |  |  |  |  |
| Healthy | — | — |  | — | — |  | — | — |  |
| Intermediate | 2.81 | 2.63, 3.00 | <0.001 | 3.01 | 2.73, 3.32 | <0.001 | 2.64 | 2.42, 2.88 | <0.001 |
| Unhealthy | 8.84 | 8.29, 9.42 | <0.001 | 9.38 | 8.55, 10.3 | <0.001 | 8.31 | 7.61, 9.06 | <0.001 |
| Smoking |  |  |  |  |  |  |  |  |  |
| Healthy | — | — |  | — | — |  | — | — |  |
| Intermediate | 1.15 | 1.11, 1.20 | <0.001 | 1.06 | 0.99, 1.12 | 0.10 | 1.23 | 1.17, 1.29 | <0.001 |
| Unhealthy | 1.65 | 1.56, 1.74 | <0.001 | 1.74 | 1.59, 1.91 | <0.001 | 1.62 | 1.51, 1.74 | <0.001 |
| Physical activity |  |  |  |  |  |  |  |  |  |
| Healthy | — | — |  | — | — |  | — | — |  |
| Intermediate | 1.03 | 0.99, 1.08 | 0.11 | 0.98 | 0.92, 1.04 | 0.50 | 1.07 | 1.02, 1.13 | 0.009 |
| Unhealthy | 1.41 | 1.33, 1.49 | <0.001 | 1.31 | 1.20, 1.43 | <0.001 | 1.48 | 1.37, 1.59 | <0.001 |
| Diet |  |  |  |  |  |  |  |  |  |
| Healthy | — | — |  | — | — |  | — | — |  |
| Unhealthy | 1.13 | 1.09, 1.17 | <0.001 | 1.09 | 1.03, 1.15 | 0.004 | 1.16 | 1.10, 1.21 | <0.001 |

*Results from Cox proportional hazards model including the multi-ancestry GRS tertiles, BMI (three levels), smoking (three levels), physical activity (three levels), and diet (two levels); models were additionally adjusted for age at enrollment, sex (sex-combined only), years in education, household income, Townsend Deprivation Index, and the first 16 genotype PC’s.*

*Abbreviations: CI, confidence interval; GRS, genetic risk score; HR, hazard ratio; T2D, type 2 diabetes*

Table S7. Independent associations of continuous GRS (multi-ancestry) and individual lifestyle factors with incident T2D risk

|  | **Sex-combined** | | | **Female** | | | **Male** | | |
| --- | --- | --- | --- | --- | --- | --- | --- | --- | --- |
| **Characteristic** | **HR** | **95% CI** | **p-value** | **HR** | **95% CI** | **p-value** | **HR** | **95% CI** | **p-value** |
| Multi-ancestry GRS | 1.51 | 1.48, 1.54 | <0.001 | 1.52 | 1.48, 1.56 | <0.001 | 1.50 | 1.47, 1.54 | <0.001 |
| BMI |  |  |  |  |  |  |  |  |  |
| Healthy | — | — |  | — | — |  | — | — |  |
| Intermediate | 2.80 | 2.62, 2.99 | <0.001 | 3.00 | 2.72, 3.30 | <0.001 | 2.63 | 2.41, 2.87 | <0.001 |
| Unhealthy | 8.80 | 8.26, 9.37 | <0.001 | 9.32 | 8.50, 10.2 | <0.001 | 8.29 | 7.59, 9.04 | <0.001 |
| Smoking |  |  |  |  |  |  |  |  |  |
| Healthy | — | — |  | — | — |  | — | — |  |
| Intermediate | 1.16 | 1.11, 1.20 | <0.001 | 1.06 | 0.99, 1.13 | 0.085 | 1.23 | 1.17, 1.29 | <0.001 |
| Unhealthy | 1.65 | 1.56, 1.74 | <0.001 | 1.75 | 1.60, 1.91 | <0.001 | 1.62 | 1.51, 1.74 | <0.001 |
| Physical activity |  |  |  |  |  |  |  |  |  |
| Healthy | — | — |  | — | — |  | — | — |  |
| Intermediate | 1.03 | 0.99, 1.08 | 0.12 | 0.98 | 0.92, 1.04 | 0.050 | 1.07 | 1.02, 1.13 | 0.010 |
| Unhealthy | 1.41 | 1.33, 1.49 | <0.001 | 1.31 | 1.20, 1.44 | <0.001 | 1.48 | 1.38, 1.59 | <0.001 |
| Diet |  |  |  |  |  |  |  |  |  |
| Healthy | — | — |  | — | — |  | — | — |  |
| Unhealthy | 1.13 | 1.08, 1.17 | <0.001 | 1.09 | 1.03, 1.15 | 0.005 | 1.16 | 1.10, 1.21 | <0.001 |

*Results from Cox proportional hazards model including the continuous multi-ancestry GRS, BMI (three levels), smoking (three levels), physical activity (three levels), and diet (two levels); models were additionally adjusted for age at enrollment, sex (sex-combined only), years in education, household income, Townsend Deprivation Index, and the first 16 genotype PC’s. The association tested was based on 1-standard deviation increase in the multi-ancestry GRS.*

*Abbreviations: CI, confidence interval; GRS, genetic risk score; HR, hazard ratio; T2D, type 2 diabetes*

**Table S8. Association of combined genetic (ancestry-specific) and lifestyle risk and T2D**

|  | | **Sex-combined** | | | **Female** | | | | **Male** | | | |
| --- | --- | --- | --- | --- | --- | --- | --- | --- | --- | --- | --- | --- |
| **GRS tertile** | **Lifestyle** | **HR** | **95% CI** | **p-value** | | **HR** | **95% CI** | **p-value** | | **HR** | **95% CI** | **p-value** |
| Low | Healthy | — | — |  | | — | — |  | | — | — |  |
|  | Intermediate | 2.55 | 2.22, 2.93 | <0.001 | | 2.59 | 2.12, 3.16 | <0.001 | | 2.50 | 2.06, 3.05 | <0.001 |
|  | Unhealthy | 7.61 | 6.37, 9.09 | <0.001 | | 7.43 | 5.63, 9.79 | <0.001 | | 7.61 | 6.00, 9.64 | <0.001 |
| Moderate | Healthy | 1.65 | 1.39, 1.95 | <0.001 | | 1.82 | 1.44, 2.29 | <0.001 | | 1.49 | 1.16, 1.90 | 0.016 |
|  | Intermediate | 3.99 | 3.48, 4.58 | <0.001 | | 3.93 | 3.24, 4.78 | <0.001 | | 3.98 | 3.28, 4.84 | <0.001 |
|  | Unhealthy | 12.73 | 10.84, 14.95 | <0.001 | | 13.52 | 10.65, 17.15 | <0.001 | | 12.20 | 9.79, 15.21 | <0.001 |
| High | Healthy | 2.84 | 2.43, 3.31 | <0.001 | | 2.92 | 2.35, 3.63 | <0.001 | | 2.76 | 2.21, 3.44 | <0.001 |
|  | Intermediate | 6.63 | 5.79, 7.59 | <0.001 | | 6.85 | 5.66, 8.30 | <0.001 | | 6.41 | 5.29, 7.77 | <0.001 |
|  | Unhealthy | 17.82 | 15.29, 20.77 | <0.001 | | 20.14 | 16.12, 25.16 | <0.001 | | 16.38 | 13.23, 20.26 | <0.001 |

*Results from Cox proportional hazards model, adjusted for age at enrollment, sex (sex-combined analysis only), years in education, household income, Townsend Deprivation Index, and the first 16 genotype PC’s.*

*Abbreviations: CI, confidence interval; HR, hazard ratio; T2D, type 2 diabetes*

Table S9. Associations of continuous GRS (multi-ancestry) and the three-level lifestyle category with incident T2D risk, including the multiplicative interaction between the GRS and lifestyle category

|  | **Sex-combined** | | | **Female** | | | **Male** | | |
| --- | --- | --- | --- | --- | --- | --- | --- | --- | --- |
| **Characteristic** | **HR** | **95% CI** | **p-value** | **HR** | **95% CI** | **p-value** | **HR** | **95% CI** | **p-value** |
| Multi-ancestry GRS | 1.58 | 1.49, 1.67 | <0.001 | 1.54 | 1.43, 1.67 | <0.001 | 1.62 | 1.49, 1.77 | <0.001 |
| Lifestyle classification |  |  |  |  |  |  |  |  |  |
| Healthy | — | — |  | — | — |  | — | — |  |
| Intermediate | 2.40 | 2.25, 2.56 | <0.001 | 2.30 | 2.10, 2.52 | <0.001 | 2.50 | 2.28, 2.75 | <0.001 |
| Unhealthy | 7.08 | 6.52, 7.70 | <0.001 | 7.01 | 6.18, 7.94 | <0.001 | 7.24 | 6.45, 8.11 | <0.001 |
| Interaction |  |  |  |  |  |  |  |  |  |
| Multi-ancestry GRS*Intermediate | 0.97 | 0.91, 1.03 | 0.30 | 1.00 | 0.92, 1.09 | 0.90 | 0.94 | 0.86, 1.03 | 0.20 |
| Multi-ancestry GRS*Unhealthy | 0.91 | 0.84, 0.98 | 0.013 | 0.97 | 0.87, 1.09 | 0.60 | 0.86 | 0.77, 0.95 | 0.004 |

*Results from Cox proportional hazards model including the continuous standardized GRS value and the three-level lifestyle classification variable and their interaction terms; model is further adjusted for age at enrollment, sex (sex-combined only), years in education, household income, Townsend Deprivation Index, and the first 16 genotype PC’s. The association tested was based on 1-standard deviation increase in the multi-ancestry GRS.*

*Abbreviations: CI, confidence interval; GRS, genetic risk score; HR, hazard ratio; T2D, type 2 diabetes*

**Table S10. Associations of GRS tertile (multi-ancestry) and the three-level lifestyle category with incident T2D risk, including the multiplicative interaction between the GRS tertiles and lifestyle category**

|  | **Sex-combined** | | | **Female** | | | **Male** | | |
| --- | --- | --- | --- | --- | --- | --- | --- | --- | --- |
| **Characteristic** | **HR** | **95% CI** | **p-value** | **HR** | **95% CI** | **p-value** | **HR** | **95% CI** | **p-value** |
| Multi-ancestry GRS tertile |  |  |  |  |  |  |  |  |  |
| Low | — | — |  | — | — |  | — | — |  |
| Moderate | 1.51 | 1.28, 1.78 | 0.001 | 1.68 | 1.34, 2.10 | 0.001 | 1.34 | 1.06, 1.71 | 0.016 |
| High | 2.68 | 2.31, 3.11 | <0.001 | 2.72 | 2.21, 3.35 | <0.001 | 2.64 | 2.13, 3.27 | <0.001 |
| Lifestyle classification |  |  |  |  |  |  |  |  |  |
| Healthy | — | — |  | — | — |  | — | — |  |
| Intermediate | 2.37 | 2.07, 2.70 | <0.001 | 2.40 | 1.99, 2.89 | <0.001 | 2.33 | 1.93, 2.81 | <0.001 |
| Unhealthy | 7.11 | 6.00, 8.43 | <0.001 | 6.85 | 5.25, 8.94 | <0.001 | 7.21 | 5.74, 9.05 | <0.001 |
| Interaction |  |  |  |  |  |  |  |  |  |
| Moderate*Intermediate | 1.05 | 0.89, 1.25 | 0.6 | 0.93 | 0.73, 1.18 | 0.6 | 1.20 | 0.93, 1.54 | 0.2 |
| High*Intermediate | 0.98 | 0.83, 1.14 | 0.8 | 0.96 | 0.77, 1.21 | 0.8 | 0.99 | 0.79, 1.24 | >0.9 |
| Moderate*Unhealthy | 1.11 | 0.90, 1.39 | 0.3 | 1.08 | 0.77, 1.50 | 0.7 | 1.20 | 0.89, 1.62 | 0.2 |
| High*Unhealthy | 0.86 | 0.70, 1.05 | 0.14 | 0.97 | 0.71, 1.33 | 0.9 | 0.80 | 0.61, 1.05 | 0.10 |

*Results from Cox proportional hazards model including the GRS tertile and the three-level lifestyle classification variable and their interaction terms; model is further adjusted for age at enrollment, sex (sex-combined only), years in education, household income, Townsend Deprivation Index, and the first 16 genotype PC’s. The association tested was based on 1-standard deviation increase in the multi-ancestry GRS.*

*Abbreviations: CI, confidence interval; GRS, genetic risk score; HR, hazard ratio; T2D, type 2 diabetes*

Table S11. Association of combined genetic (multi-ancestry) and lifestyle risk and T2D stratified by ancestry

|  |  | **AFR** | | | **EAS** | | | **EUR** | | | **SAS** | | |
| --- | --- | --- | --- | --- | --- | --- | --- | --- | --- | --- | --- | --- | --- |
| **GRS tertile** | **Lifestyle** | **HR** | **95% CI** | **p-value** | **HR** | **95% CI** | **p-value** | **HR** | **95% CI** | **p-value** | **HR** | **95% CI** | **p-value** |
| Low | |  |  |  |  |  |  |  |  |  |  |  |  |
|  | Healthy | — | — |  | — | — |  | — | — |  | — | — |  |
|  | Intermediate | 2.42 | 1.22, 4.80 | 0.01 | 2.03 | 0.67, 6.12 | 0.208 | 2.66 | 2.28, 3.09 | <0.001 | 1.25 | 0.85, 1.84 | 0.26 |
|  | Unhealthy | 2.17 | 0.77, 6.11 | 0.14 | 0.00 | 0.00, Inf | 0.997 | 8.32 | 6.89, 10.05 | <0.001 | 4.46 | 2.43, 8.18 | <0.001 |
| Moderate | |  |  |  |  |  |  |  |  |  |  |  |  |
|  | Healthy | 0.84 | 0.32, 2.18 | 0.72 | 0.98 | 0.18, 5.42 | 0.982 | 1.66 | 1.38, 2.00 | <0.001 | 1.17 | 0.71, 1.92 | 0.534 |
|  | Intermediate | 2.76 | 1.40, 5.45 | 0.003 | 3.04 | 0.99, 9.33 | 0.052 | 4.28 | 3.69, 4.97 | <0.001 | 1.94 | 1.32, 2.84 | <0.001 |
|  | Unhealthy | 5.41 | 2.27, 12.91 | <0.001 | 0.00 | 0.00, Inf | 0.998 | 13.99 | 11.78, 16.61 | <0.001 | 7.99 | 4.40, 14.51 | <0.001 |
| High | |  |  |  |  |  |  |  |  |  |  |  |  |
|  | Healthy | 2.49 | 1.14, 5.42 | 0.02 | 3.94 | 0.94, 16.48 | 0.061 | 3.00 | 2.53, 3.55 | <0.001 | 1.51 | 0.93, 2.45 | 0.09 |
|  | Intermediate | 3.19 | 1.62, 6.28 | <0.001 | 4.77 | 1.46, 15.61 | 0.001 | 7.20 | 6.21, 8.34 | <0.001 | 2.91 | 1.99, 4.25 | <0.001 |
|  | Unhealthy | 5.12 | 2.20, 11.91 | <0.001 | 17.88 | 1.78, 179.97 | 0.014 | 19.56 | 16.6, 23.07 | <0.001 | 5.60 | 2.73, 11.46 | <0.001 |

*Results from Cox proportional hazards model, adjusted for age at enrollment, sex, years in education, household income, Townsend
Deprivation Index, and the first 16 genotype PC’s.*

*Abbreviations: AFR, African; CI, confidence interval; EAS, East Asian; EUR, European; HR, hazard ratio; SAS South Asian; T2D, type 2 diabetes*

**Table S12. Independent associations of GRS tertile (ancestry-specific) and individual lifestyle factors with incident T2D risk**

|  | **Sex-combined** | | | **Female** | | | **Male** | | |
| --- | --- | --- | --- | --- | --- | --- | --- | --- | --- |
| **Characteristic** | **HR** | **95% CI** | **p-value** | **HR** | **95% CI** | **p-value** | **HR** | **95% CI** | **p-value** |
| Ancestry-specific GRS tertile |  |  |  |  |  |  |  |  |  |
| Low | — | — |  | — | — |  | — | — |  |
| Moderate | 1.56 | 1.48, 1.64 | <0.001 | 1.56 | 1.43, 1.69 | <0.001 | 1.57 | 1.47, 1.67 | <0.001 |
| High | 2.51 | 2.39, 2.63 | <0.001 | 2.59 | 2.40, 2.80 | <0.001 | 2.46 | 2.31, 2.61 | <0.001 |
| BMI |  |  |  |  |  |  |  |  |  |
| Healthy | — | — |  | — | — |  | — | — |  |
| Intermediate | 2.81 | 2.63, 3.00 | <0.001 | 3.01 | 2.73, 3.33 | <0.001 | 2.63 | 2.41, 2.88 | <0.001 |
| Unhealthy | 8.85 | 8.30, 9.44 | <0.001 | 9.42 | 8.58, 10.36 | <0.001 | 8.30 | 7.59, 9.06 | <0.001 |
| Smoking |  |  |  |  |  |  |  |  |  |
| Healthy | — | — |  | — | — |  | — | — |  |
| Intermediate | 1.16 | 1.11, 1.20 | <0.001 | 1.05 | 0.98, 1.12 | 0.158 | 1.23 | 1.17, 1.30 | <0.001 |
| Unhealthy | 1.66 | 1.57, 1.76 | <0.001 | 1.78 | 1.63, 1.95 | <0.001 | 1.62 | 1.51, 1.74 | <0.001 |
| Physical activity |  |  |  |  |  |  |  |  |  |
| Healthy | — | — |  | — | — |  | — | — |  |
| Intermediate | 1.03 | 0.99, 1.08 | 0.119 | 0.97 | 0.91, 1.04 | 0.385 | 1.08 | 1.02, 1.14 | 0.007 |
| Unhealthy | 1.41 | 1.33, 1.49 | <0.001 | 1.31 | 1.20, 1.44 | <0.001 | 1.48 | 1.37, 1.59 | <0.001 |
| Diet |  |  |  |  |  |  |  |  |  |
| Healthy | — | — |  | — | — |  | — | — |  |
| Unhealthy | 1.12 | 1.08, 1.17 | <0.001 | 1.09 | 1.03, 1.15 | 0.004 | 1.15 | 1.10, 1.21 | <0.001 |

*Results from Cox proportional hazards model including the ancestry-specific GRS tertiles, BMI (three levels), smoking (three levels), physical activity (three levels), and diet (two levels); models were additionally adjusted for age at enrollment, sex (sex-combined only), years in education, household income, Townsend Deprivation Index, and the first 16 genotype PC’s.*

*Abbreviations: CI, confidence interval; GRS, genetic risk score; HR, hazard ratio; T2D, type 2 diabetes*

Table S13. Independent associations of GRS (ancestry-specific) and individual lifestyle factors with incident T2D risk

|  | **Sex-combined** | | | **Female** | | | **Male** | | |
| --- | --- | --- | --- | --- | --- | --- | --- | --- | --- |
| **Characteristic** | **HR** | **95% CI** | **p-value** | **HR** | **95% CI** | **p-value** | **HR** | **95% CI** | **p-value** |
| Ancestry-specific GRS (standardized) | 1.51 | 1.48, 1.54 | <0.001 | 1.52 | 1.48, 1.57 | <0.001 | 1.50 | 1.47, 1.54 | <0.001 |
| BMI |  |  |  |  |  |  |  |  |  |
| Healthy | — | — |  | — | — |  | — | — |  |
| Intermediate | 2.79 | 2.61, 2.98 | <0.001 | 3.01 | 2.72, 3.32 | <0.001 | 2.62 | 2.39, 2.86 | <0.001 |
| Unhealthy | 8.79 | 8.25, 9.38 | <0.001 | 9.36 | 8.52, 10.29 | <0.001 | 8.25 | 7.56, 9.02 | <0.001 |
| Smoking |  |  |  |  |  |  |  |  |  |
| Healthy | — | — |  | — | — |  | — | — |  |
| Intermediate | 1.16 | 1.11, 1.20 | <0.001 | 1.05 | 0.98, 1.12 | 0.140 | 1.23 | 1.17, 1.30 | <0.001 |
| Unhealthy | 1.67 | 1.58, 1.76 | <0.001 | 1.78 | 1.63, 1.95 | <0.001 | 1.63 | 1.52, 1.75 | <0.001 |
| Physical activity |  |  |  |  |  |  |  |  |  |
| Healthy | — | — |  | — | — |  | — | — |  |
| Intermediate | 1.03 | 0.99, 1.08 | 0.125 | 0.97 | 0.91, 1.04 | 0.389 | 1.08 | 1.02, 1.14 | 0.008 |
| Unhealthy | 1.42 | 1.34, 1.50 | <0.001 | 1.32 | 1.20, 1.44 | <0.001 | 1.48 | 1.38, 1.60 | <0.001 |
| Diet |  |  |  |  |  |  |  |  |  |
| Healthy | — | — |  | — | — |  | — | — |  |
| Unhealthy | 1.12 | 1.08, 1.17 | <0.001 | 1.09 | 1.03, 1.15 | 0.004 | 1.15 | 1.10, 1.21 | <0.001 |

*Results from Cox proportional hazards model including the continuous ancestry-specific GRS, BMI (three levels), smoking (three levels), physical activity (three levels), and diet (two levels); models were additionally adjusted for age at enrollment, sex (sex-combined only), years in education, household income, Townsend Deprivation Index, and the first 16 genotype PC’s. The association tested was based on 1-standard deviation increase in the ancestry-specific GRS.*

*Abbreviations: CI, confidence interval; GRS, genetic risk score; HR, hazard ratio; T2D, type 2 diabetes*

Table S14. Independent associations of GRS tertiles (ancestry-specific) and the three-level lifestyle category with incident T2D risk

|  | **Sex-combined** | | | | **Female** | | | **Male** | | |
| --- | --- | --- | --- | --- | --- | --- | --- | --- | --- | --- |
| **Characteristic** | **HR** | | **95% CI** | **p-value** | **HR** | **95% CI** | **p-value** | **HR** | **95% CI** | **p-value** |
| Ancestry-specific GRS | |  |  |  |  |  |  |  |  |  |
| Low | | — | — |  | — | — |  | — | — |  |
| Moderate | | 1.58 | 1.51, 1.67 | <0.001 | 1.58 | 1.46, 1.71 | <0.001 | 1.59 | 1.49, 1.69 | <0.001 |
| High | | 2.59 | 2.47, 2.71 | <0.001 | 2.68 | 2.49, 2.89 | <0.001 | 2.52 | 2.38, 2.68 | <0.001 |
| Lifestyle classification | |  |  |  |  |  |  |  |  |  |
| Healthy | | — | — |  | — | — |  | — | — |  |
| Intermediate | | 2.40 | 2.26, 2.56 | <0.001 | 2.33 | 2.14, 2.54 | <0.001 | 2.46 | 2.26, 2.69 | <0.001 |
| Unhealthy | | 6.96 | 6.43, 7.53 | <0.001 | 7.17 | 6.37, 8.07 | <0.001 | 6.91 | 6.20, 7.70 | <0.001 |
| *Results from Cox proportional hazards model including the ancestry-specific GRS tertiles and the three-level lifestyle classification variable; model is further adjusted for age at enrollment, sex (sex-combined only), years in education, household income, Townsend Deprivation Index, and the first 16 genotype PC’s.*  *Abbreviations: CI, confidence interval; GRS, genetic risk score; HR, hazard ratio; T2D, type 2 diabetes* | | | | | | | | | | |

Table S15. Independent associations of continuous GRS (ancestry-specific) and the three-level lifestyle category with incident T2D risk

|  | | **Sex-combined** | | | **Female** | | | **Male** | | |
| --- | --- | --- | --- | --- | --- | --- | --- | --- | --- | --- |
| **Characteristic** | **HR** | | **95% CI** | **p-value** | **HR** | **95% CI** | **p-value** | **HR** | **95% CI** | **p-value** |
| Ancestry-specific GRS (standardized) | 1.53 | | 1.50, 1.56 | <0.001 | 1.55 | 1.50, 1.59 | <0.001 | 1.52 | 1.48, 1.55 | <0.001 |
| Lifestyle classification |  | |  |  |  |  |  |  |  |  |
| Healthy | — | | — |  | — | — |  | — | — |  |
| Intermediate | 2.40 | | 2.26, 2.55 | <0.001 | 2.33 | 2.14, 2.54 | <0.001 | 2.46 | 2.25, 2.69 | <0.001 |
| Unhealthy | 6.96 | | 6.43, 7.53 | <0.001 | 7.15 | 6.35, 8.05 | <0.001 | 6.91 | 6.20, 7.70 | <0.001 |
| *Results from Cox proportional hazards model including the ancestry-specific GRS and the three-level lifestyle classification variable; model is further adjusted for age at enrollment, sex (sex-combined only), years in education, household income, Townsend Deprivation Index, and the first 16 genotype PC’s. The association tested was based on 1-standard deviation increase in the ancestry-specific GRS.*  *Abbreviations: CI, confidence interval; GRS, genetic risk score; HR, hazard ratio; T2D, type 2 diabetes* | | | | | | | | | | |

**Table S16. Association of combined genetic (ancestry-specific) and lifestyle risk and T2D stratified by ancestry**

|  |  | **Overall** | | | **AFR** | | | **EAS** | | | **EUR** | | | **SAS** | | |
| --- | --- | --- | --- | --- | --- | --- | --- | --- | --- | --- | --- | --- | --- | --- | --- | --- |
| **GRS tertile** | **Lifestyle** | **HR** | **95% CI** | **p-value** | **HR** | **95% CI** | **p-value** | **HR** | **95% CI** | **p-value** | **HR** | **95% CI** | **p-value** | **HR** | **95% CI** | **p-value** |
| Low | Healthy | — | — |  | — | — |  | — | — |  | — | — |  | — | — |  |
|  | Intermediate | 2.55 | 2.22, 2.93 | <0.001 | 2.75 | 1.34, 5.67 | 0.006 | 5.12 | 0.65, 40.60 | 0.122 | 2.67 | 2.30, 3.11 | <0.001 | 1.48 | 0.95, 2.32 | 0.083 |
|  | Unhealthy | 7.61 | 6.37, 9.09 | <0.001 | 2.53 | 0.88, 7.31 | 0.086 | 0.00 | 0.00, Inf | 0.997 | 8.24 | 6.83, 9.95 | <0.001 | 5.76 | 2.85, 11.62 | <0.001 |
| Moderate | Healthy | 1.65 | 1.39, 1.95 | <0.001 | 1.68 | 0.70, 4.01 | 0.242 | 3.48 | 0.36, 34.04 | 0.284 | 1.67 | 1.39, 2.00 | <0.001 | 1.41 | 0.83, 2.39 | 0.200 |
|  | Intermediate | 3.99 | 3.48, 4.58 | <0.001 | 3.27 | 1.60, 6.71 | 0.001 | 5.32 | 0.69, 41.25 | 0.110 | 4.26 | 3.67, 4.94 | <0.001 | 2.07 | 1.34, 3.20 | 0.001 |
|  | Unhealthy | 12.73 | 10.84, 14.95 | <0.001 | 6.37 | 2.59, 15.71 | <0.001 | 0.00 | 0.00, Inf | 0.998 | 13.94 | 11.74, 16.55 | <0.001 | 6.05 | 3.22, 11.35 | <0.001 |
| High | Healthy | 2.84 | 2.43, 3.31 | <0.001 | 2.37 | 1.02, 5.51 | 0.045 | 6.90 | 0.82, 58.23 | 0.076 | 3.00 | 2.53, 3.54 | <0.001 | 1.58 | 0.95, 2.63 | 0.078 |
|  | Intermediate | 6.63 | 5.79, 7.59 | <0.001 | 3.72 | 1.82, 7.60 | <0.001 | 11.14 | 1.48, 84.03 | 0.019 | 7.22 | 6.23, 8.36 | <0.001 | 2.82 | 1.83, 4.33 | <0.001 |
|  | Unhealthy | 17.82 | 15.29, 20.77 | <0.001 | 5.86 | 2.44, 14.05 | <0.001 | 38.66 | 2.22, 672.34 | 0.012 | 19.70 | 16.72, 23.20 | <0.001 | 7.35 | 3.76, 14.37 | <0.001 |

*Results from Cox proportional hazards model, adjusted for age at enrollment, sex, years in education, household income, Townsend Deprivation Index, and the first 16 genotype PC’s.*

*Abbreviations: AFR, African; CI, confidence interval; EAS, East Asian; EUR, European; HR, hazard ratio; SAS South Asian; T2D, type 2 diabetes*

Table S17. Population attributable fraction

|  | **Non-healthy to healthy lifestyle** | | | **Intermediate to healthy lifestyle** | | | **Unhealthy to healthy lifestyle** | | |
| --- | --- | --- | --- | --- | --- | --- | --- | --- | --- |
| **Year** | **PAF** | **CI** | **P** | **PAF** | **CI** | **P** | **PAF** | **CI** | **P** |
| 1 | 0.555 | 0.531, 0.579 | <0.001 | 0.520 | 0.494, 0.546 | <0.001 | 0.495 | 0.471, 0.518 | <0.001 |
| 2 | 0.555 | 0.53, 0.579 | <0.001 | 0.520 | 0.494, 0.546 | <0.001 | 0.493 | 0.47, 0.517 | <0.001 |
| 3 | 0.554 | 0.53, 0.578 | <0.001 | 0.519 | 0.493, 0.545 | <0.001 | 0.492 | 0.469, 0.515 | <0.001 |
| 4 | 0.554 | 0.529, 0.578 | <0.001 | 0.519 | 0.493, 0.545 | <0.001 | 0.49 | 0.467, 0.514 | <0.001 |
| 5 | 0.553 | 0.529, 0.577 | <0.001 | 0.518 | 0.492, 0.544 | <0.001 | 0.488 | 0.465, 0.512 | <0.001 |
| 6 | 0.552 | 0.528, 0.576 | <0.001 | 0.517 | 0.491, 0.543 | <0.001 | 0.487 | 0.463, 0.51 | <0.001 |
| 7 | 0.551 | 0.527, 0.575 | <0.001 | 0.517 | 0.491, 0.543 | <0.001 | 0.484 | 0.461, 0.507 | <0.001 |
| 8 | 0.55 | 0.526, 0.574 | <0.001 | 0.516 | 0.49, 0.542 | <0.001 | 0.482 | 0.459, 0.505 | <0.001 |
| 9 | 0.549 | 0.525, 0.574 | <0.001 | 0.515 | 0.489, 0.541 | <0.001 | 0.479 | 0.457, 0.502 | <0.001 |
| 10 | 0.548 | 0.524, 0.573 | <0.001 | 0.514 | 0.488, 0.54 | <0.001 | 0.477 | 0.454, 0.5 | <0.001 |
| 11 | 0.547 | 0.523, 0.572 | <0.001 | 0.513 | 0.487, 0.539 | <0.001 | 0.475 | 0.452, 0.498 | <0.001 |
| 12 | 0.546 | 0.522, 0.571 | <0.001 | 0.512 | 0.486, 0.538 | <0.001 | 0.473 | 0.45, 0.495 | <0.001 |
| 13 | 0.545 | 0.521, 0.57 | <0.001 | 0.511 | 0.485, 0.537 | <0.001 | 0.47 | 0.448, 0.493 | <0.001 |
| 14 | 0.544 | 0.52, 0.569 | <0.001 | 0.510 | 0.484, 0.536 | <0.001 | 0.468 | 0.445, 0.49 | <0.001 |
| 15 | 0.543 | 0.519, 0.568 | <0.001 | 0.509 | 0.483, 0.535 | <0.001 | 0.466 | 0.444, 0.488 | <0.001 |

*Abbreviations: PAF, Population Attributable Fraction; CI, Confidence Interval*

*Results were based on Cox proportional hazards model, adjusted for age at enrollment, sex, years in education, household income, Townsend Deprivation Index, and the first 16 genotype PC’s.*

Figure S1. Survival curves for multi-ancestry GR category

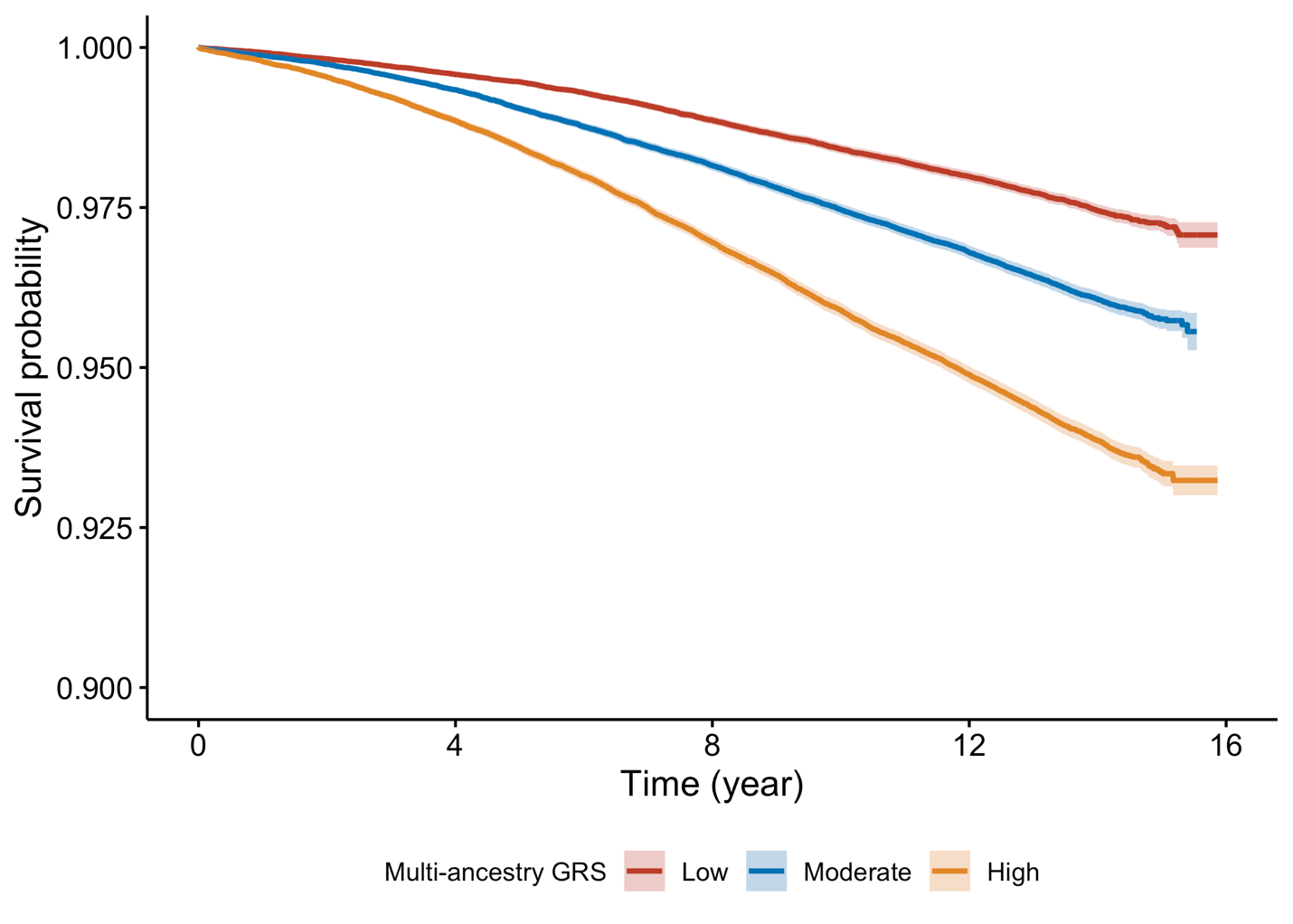

Figure S2. Survival curves for ancestry-specific GR category

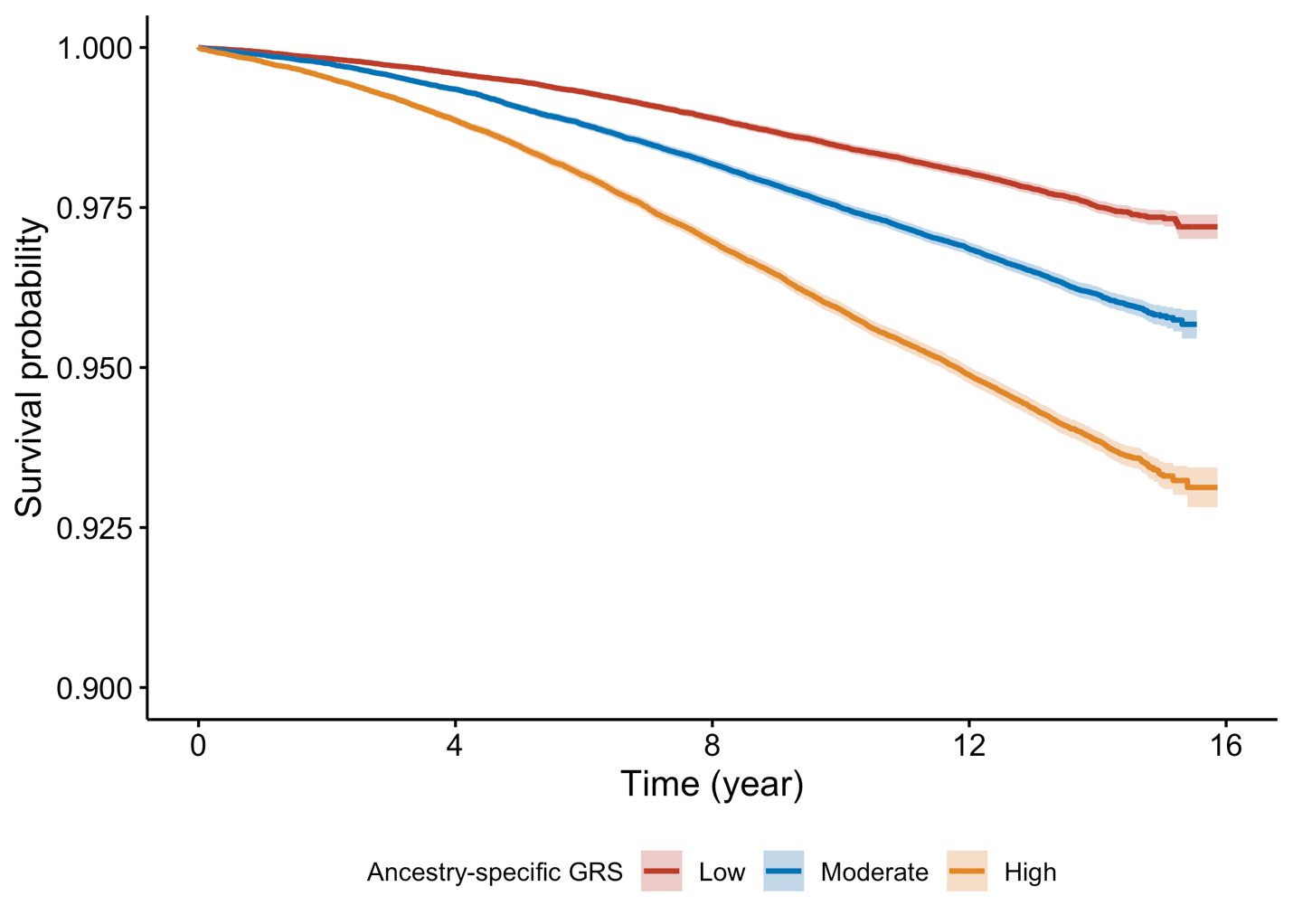

Figure S3. Survival curves for lifestyle classification

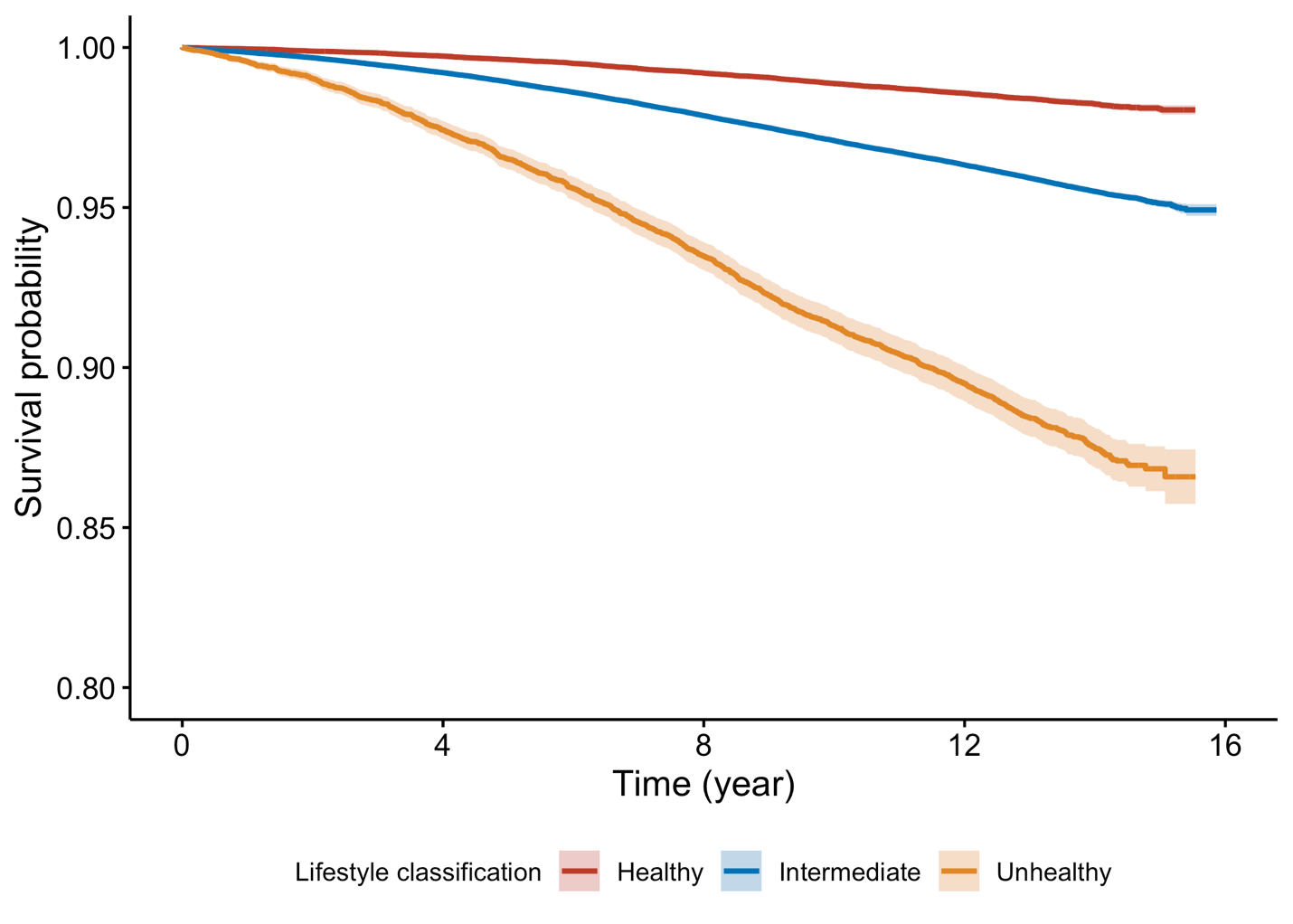

Figure S4. Survival curves for combined genetic (multi-ancestry) and lifestyle risk

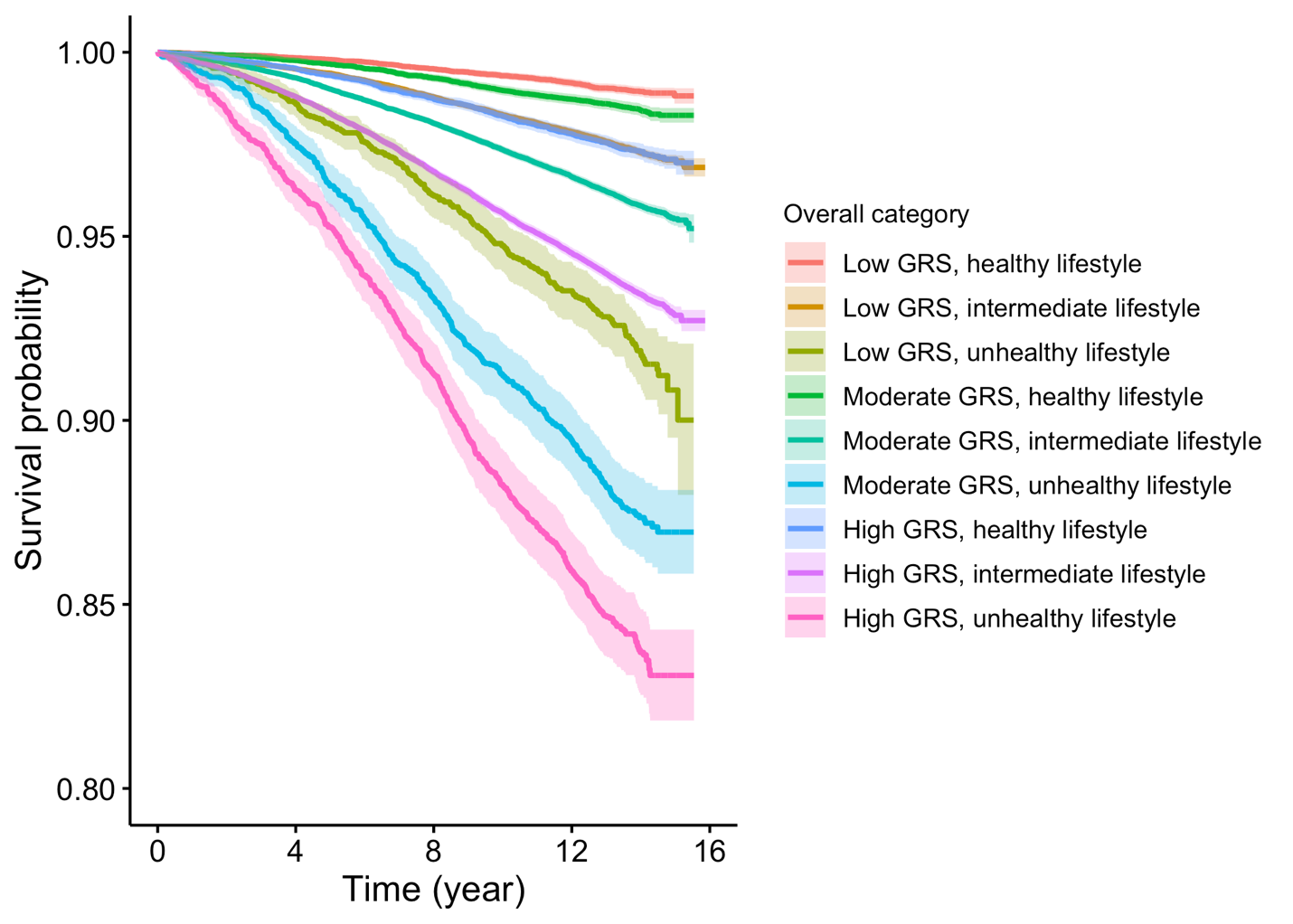

Figure S5. Survival curves for combined genetic (ancestry-specific) and lifestyle risk
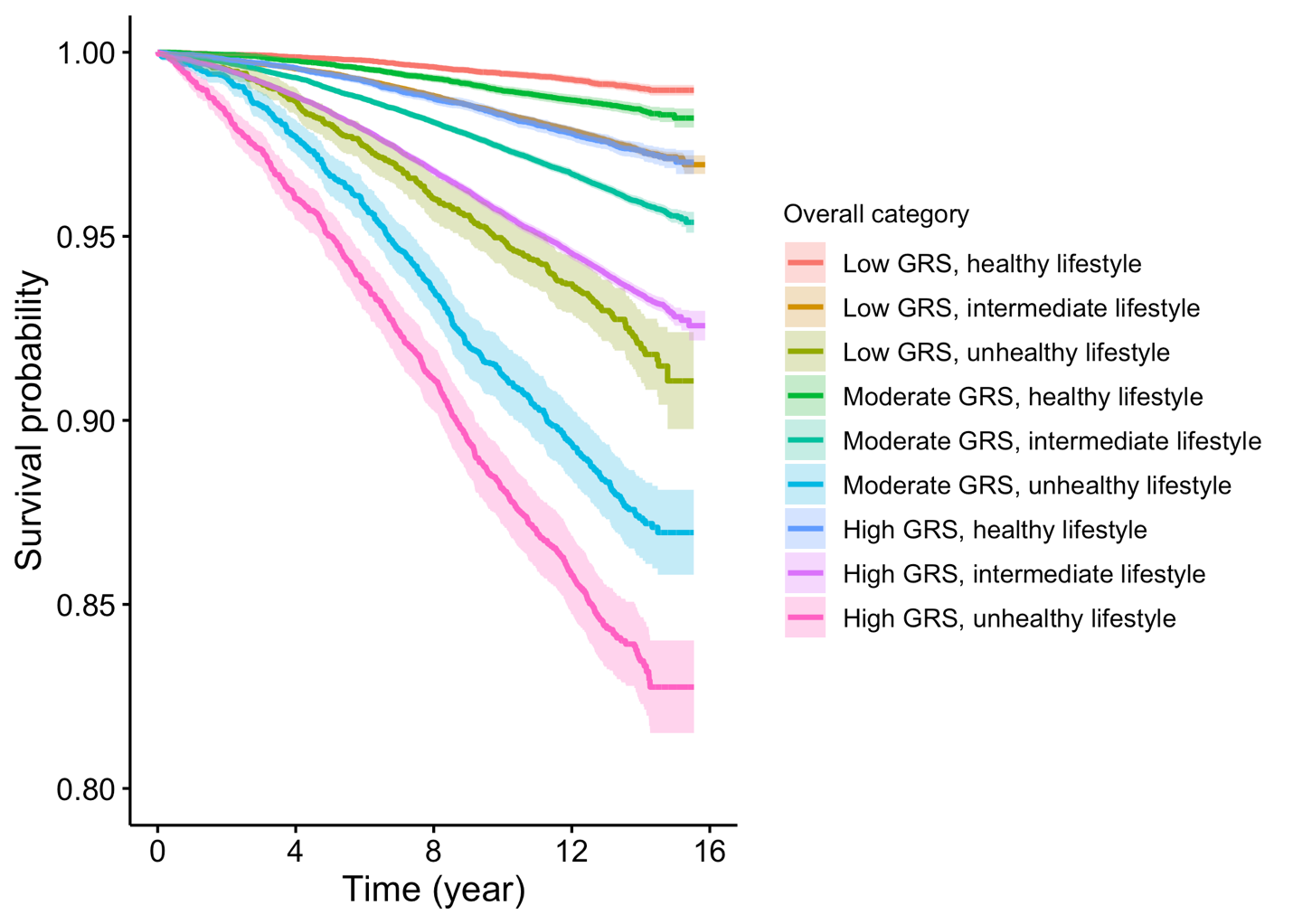

Figure S6. Graphical test of proportional hazard (PH) assumption for multi-ancestry GRS

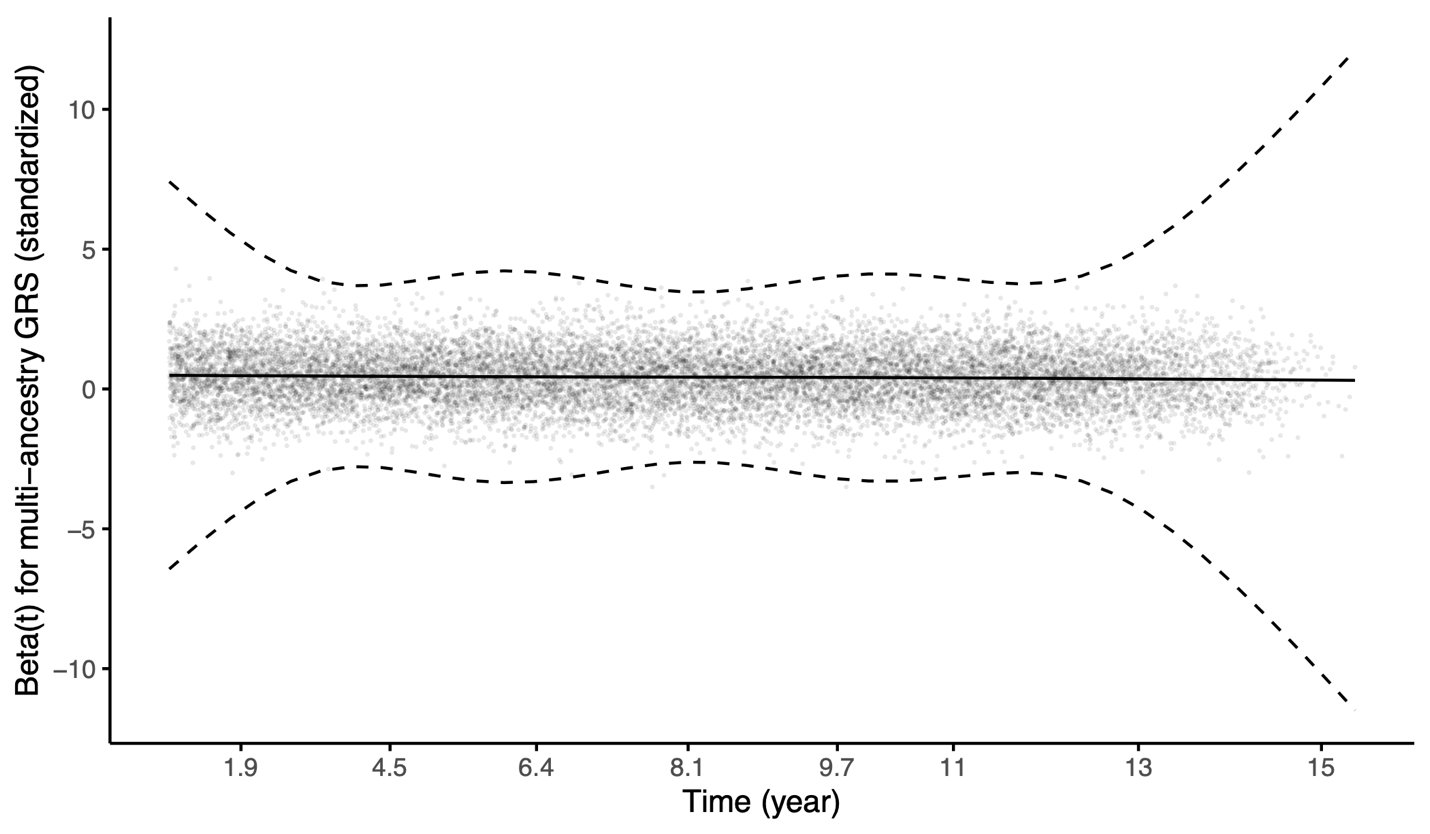

*Results from Cox proportional hazards model, adjusted for lifestyle classification, age at enrollment, sex, years in education, household income, Townsend Deprivation Index, and the first 16 genotype PC’s. The smooth line shown in the figure indicated the PH assumption was not violated.*

Figure S7. Graphical test of PH assumption for multi-ancestry GR

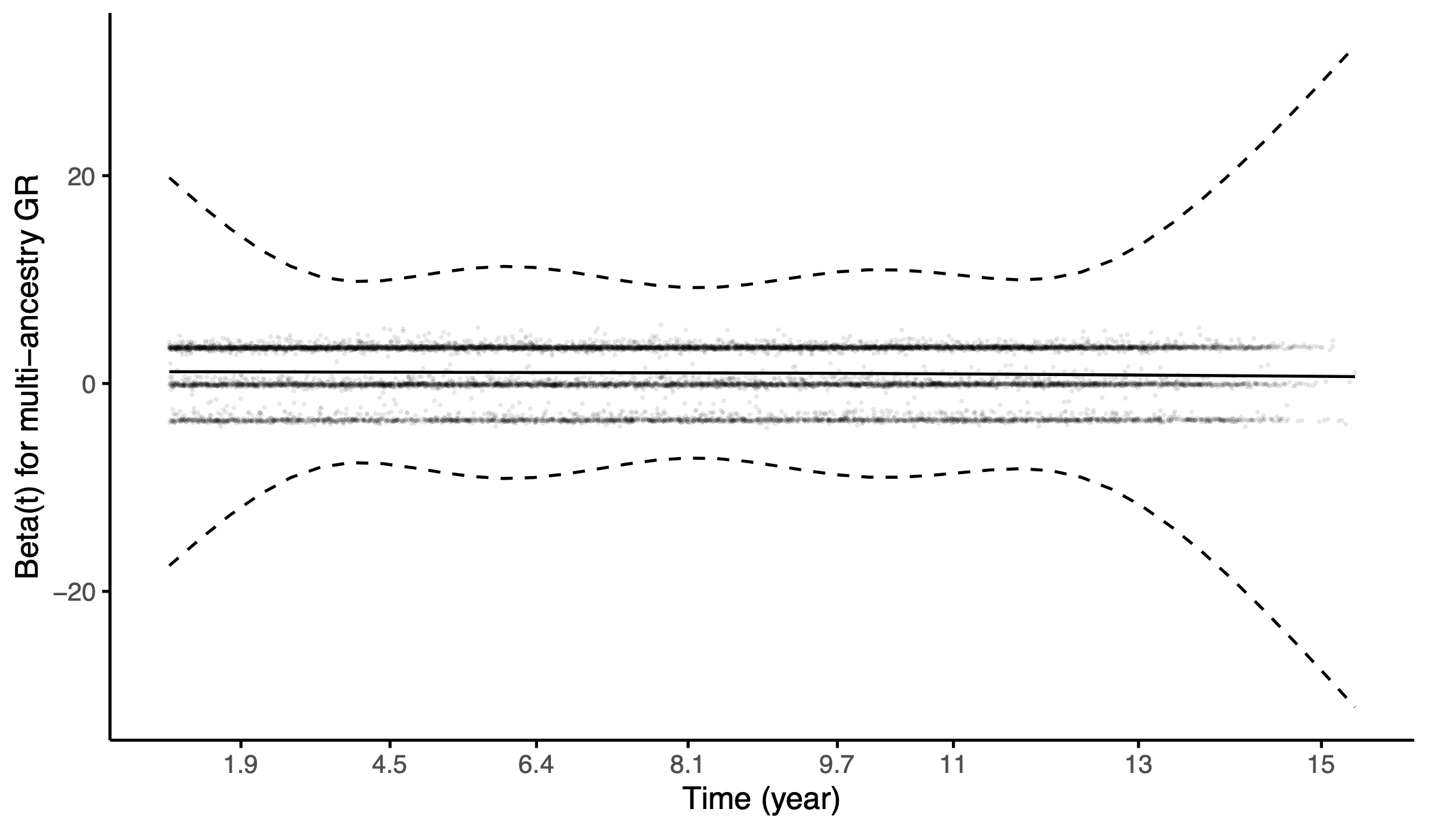

*Results from Cox proportional hazards model, adjusted for lifestyle classification, age at enrollment, sex, years in education, household income, Townsend Deprivation Index, and the first 16 genotype PC’s. The smooth line shown in the figure indicated the PH assumption was not violated.*

Figure S8. Graphical test of PH assumption for lifestyle classification

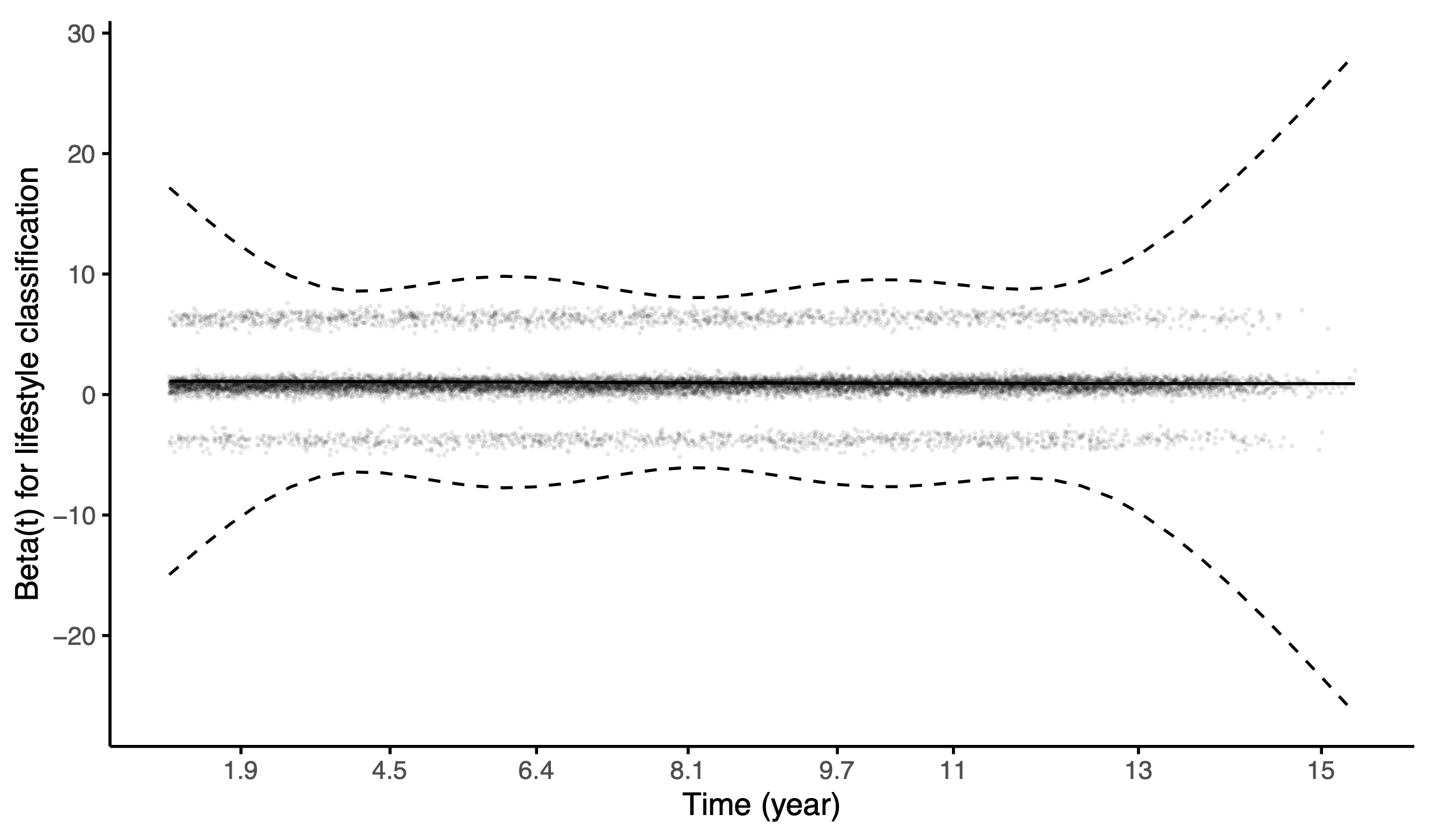

*Results from Cox proportional hazards model, adjusted for multi-ancestry GRS, age at enrollment, sex, years in education, household income, Townsend Deprivation Index, and the first 16 genotype PC’s. The smooth lines shown in the figure indicated the PH assumption was not violated.*

Figure S9. Graphical test of PH assumption for combined genetic (multi-ancestry) and lifestyle

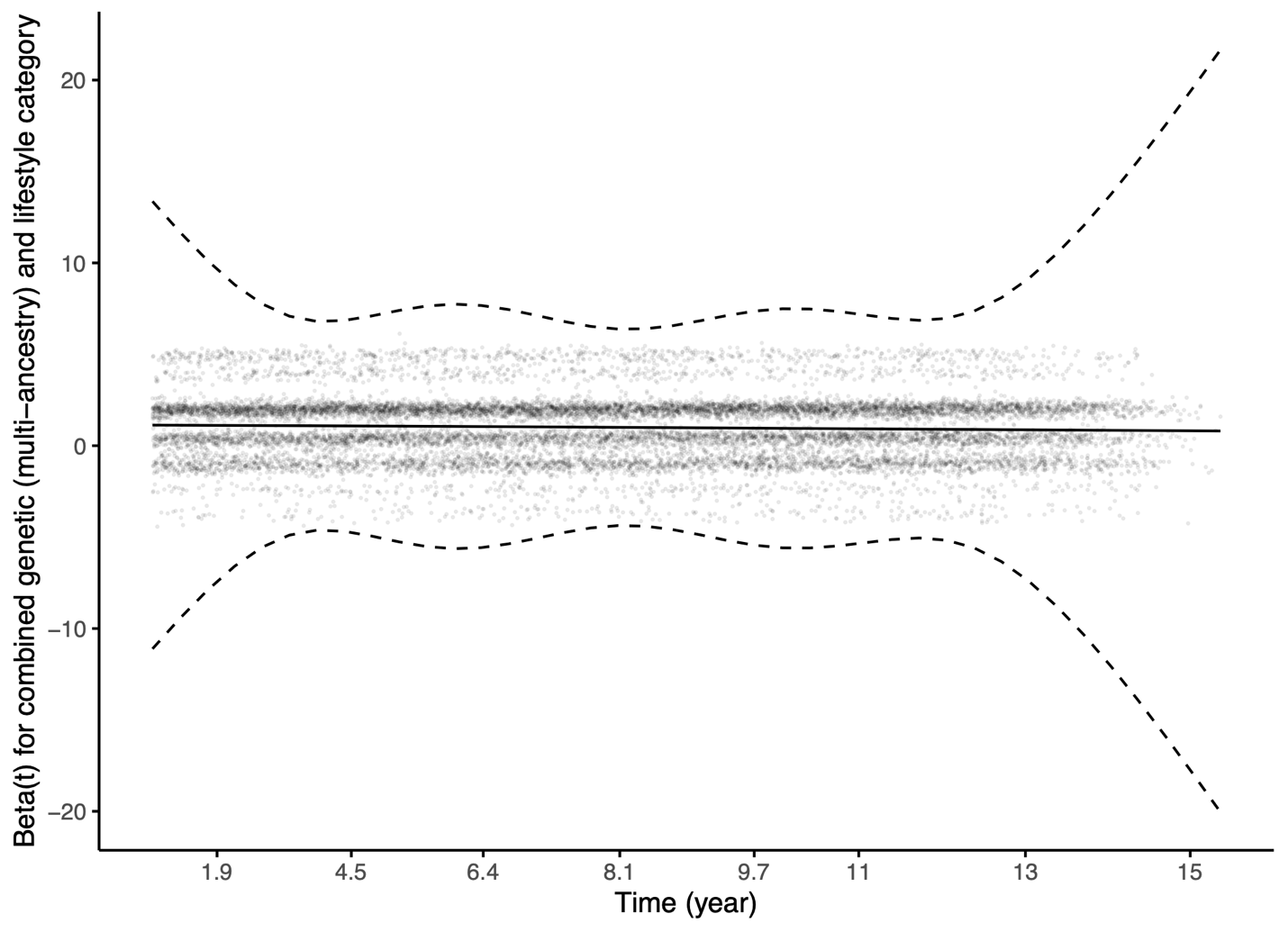

*Results from Cox proportional hazards model, adjusted for age at enrollment, sex, years in education, household income, Townsend Deprivation Index, and the first 16 genotype PC’s. The smooth line shown in the figure indicated the PH assumption was not violated.*

Figure S10. Graphical test of PH assumption for combined genetic (ancestry-specific) and lifestyle risk

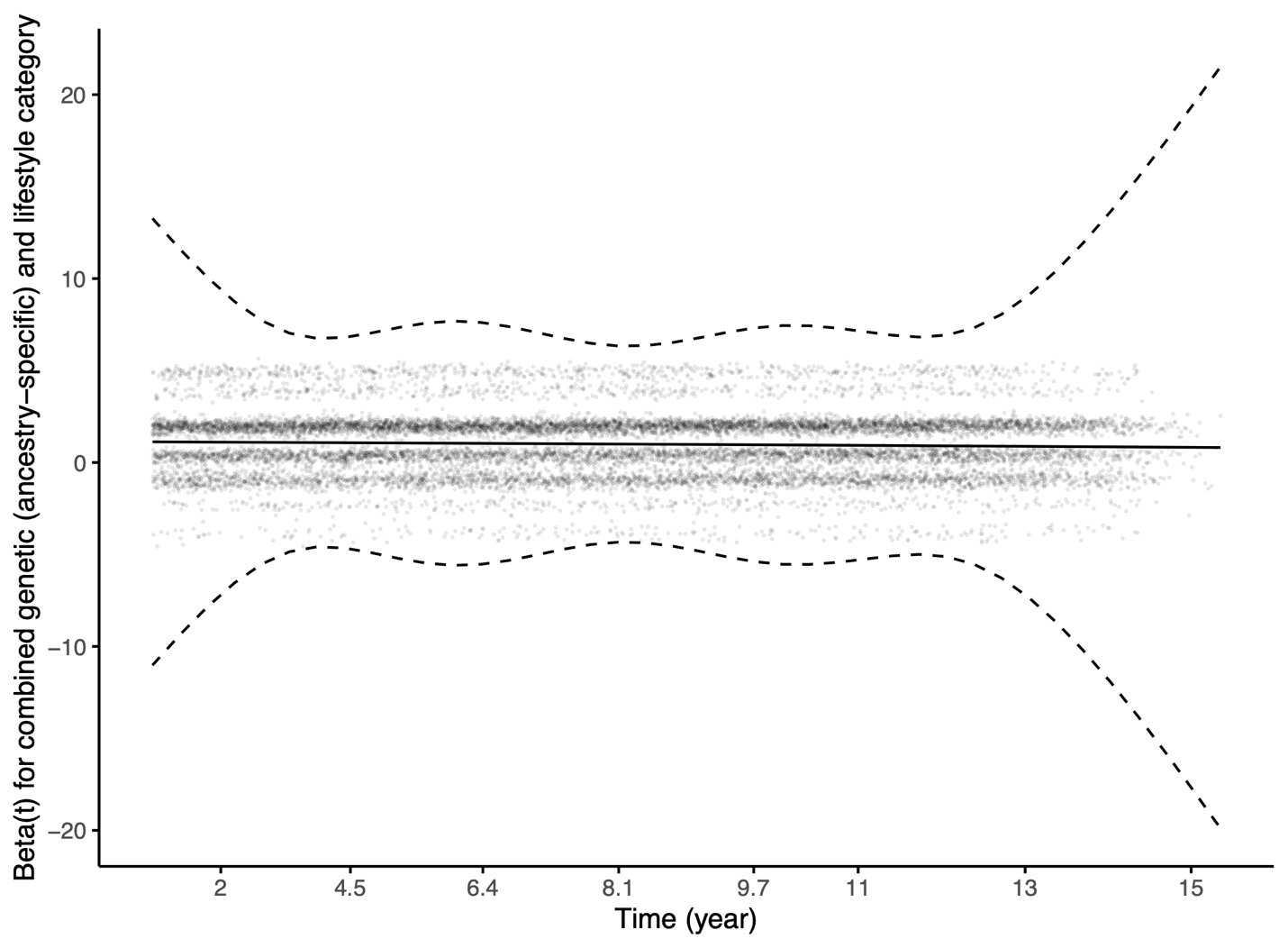

*Results from Cox proportional hazards model, adjusted for age at enrollment, sex, years in education, household income, Townsend Deprivation Index, and the first 16 genotype PC’s. The smooth line shown in the figure indicated the PH assumption was not violated.*

Figure S11. Multi-ancestry GRS by T2D

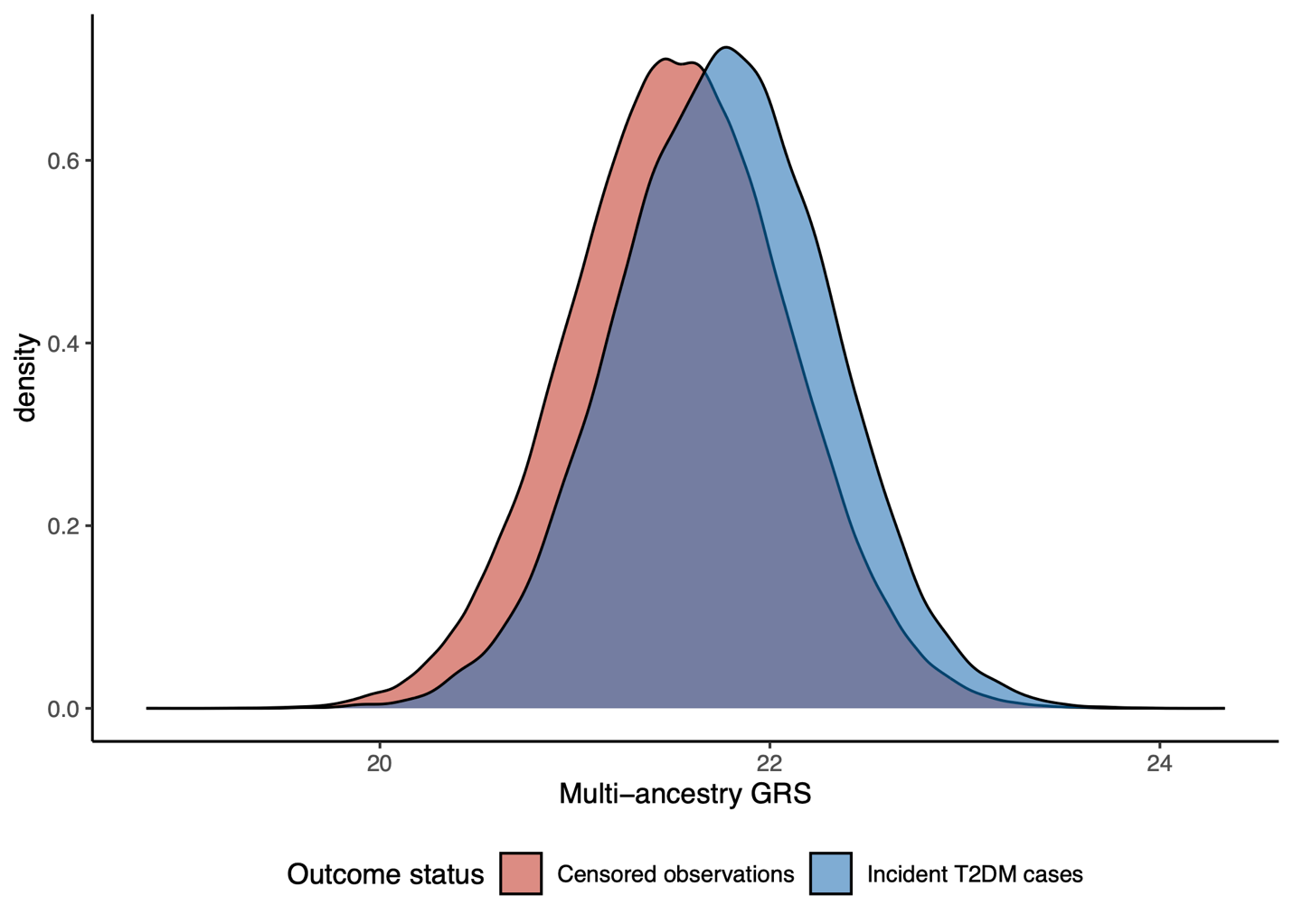

Figure S12. Ancestry-specific GRS by T2D

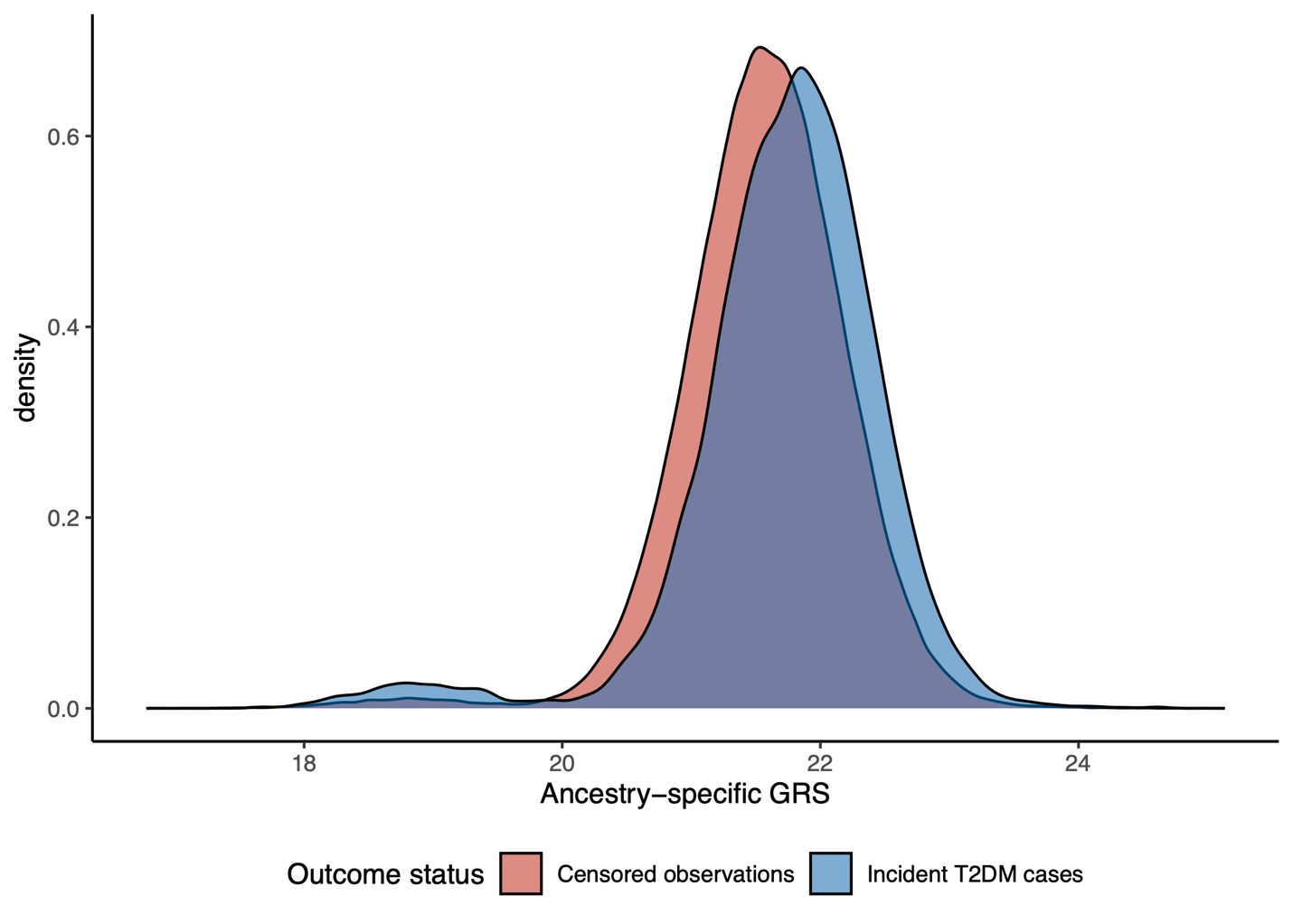

Figure S13. Correlation between multi-ancestry and ancestry-specific GRS

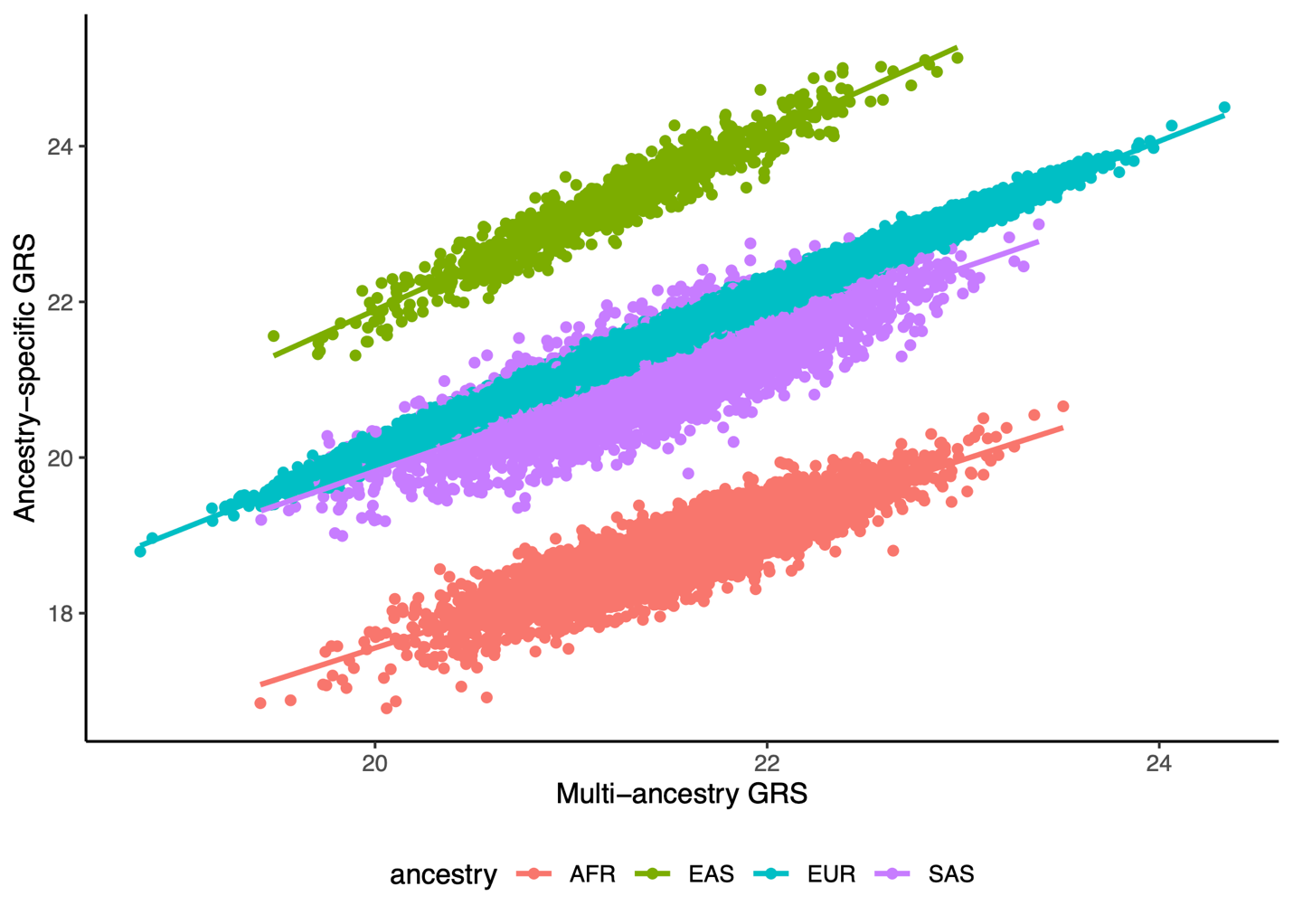

Figure S14. Association of combined genetic (ancestry-specific) and lifestyle risk with T2D

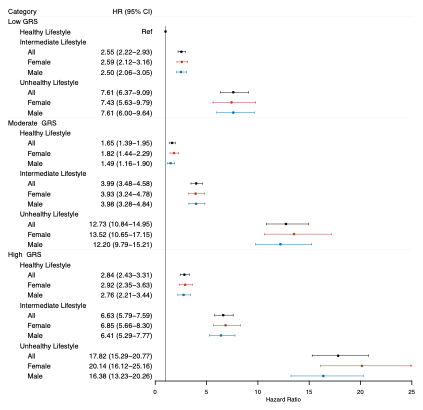

*Results from Cox proportional hazards model, adjusted for age at enrollment, sex (sex-combined only), years in education, household income, Townsend Deprivation Index, and the first 16 genotype PC’s.*
